# Patient Clusters and Cost Trajectories in Atrial Fibrillation: Evidence from the Swiss Atrial Fibrillation Cohort

**DOI:** 10.1101/2022.06.07.22275906

**Authors:** Aebersold Helena, Serra-Burriel Miquel, Foster-Witassek Fabienne, Moschovitis Giorgio, Aeschbacher Stefanie, Auricchio Angelo, Jürg H. Beer, Blozik Eva, Leo H. Bonati, Conen David, Felder Stefan, Carola A. Huber, Kühne Michael, Müller Andreas, Oberle Jolanda, Rebecca E. Paladini, Reichlin Tobias, Rodondi Nicolas, Springer Anne, Stauber Annina, Sticherling Christian, Szucs Thomas, Osswald Stefan, Schwenkglenks Matthias, the Swiss-AF Investigators

**Affiliations:** Epidemiology, Biostatistics and Prevention Institute, University of Zurich, Zurich, Switzerland; Division of Cardiology, Ente Ospedaliero Cantonale (EOC), Ospedale Regionale di Lugano, Lugano, Switzerland; Cardiology Division, Department of Medicine, University Hospital Basel, Switzerland; Cardiovascular Research Institute Basel, University Hospital Basel, Switzerland; Department of Cardiology, Instituto Cardiocentro Ticino, Ente Ospedaliero Cantonale (EOC), Lugano, Switzerland; Department of Medicine, Cantonal Hospital of Baden, Switzerland; Center for Molecular Cardiology, University of Zürich, Switzerland; Institute of Primary Care, University of Zurich, Zurich, Switzerland; Department of Neurology, University Hospital Basel, Basel Switzerland; Research Department, Reha Rheinfelden, Rheinfelden, Switzerland; Population Health Research Institute, McMaster University, Hamilton, Canada; University of Basel, Faculty of Business and Economics, Switzerland; Department of Health Sciences, Helsana Group, Zurich, Switzerland; Department of Cardiology, Triemli Hospital Zurich, Switzerland; Institute of Primary Health Care (BIHAM), University of Bern, Switzerland; Department of General Internal Medicine, Inselspital, Bern University Hospital, University of Bern, Switzerland; Department of Cardiology, Inselspital, Bern University Hospital, University of Bern, Switzerland; Institute of Pharmaceutical Medicine (ECPM), University of Basel, Basel, Switzerland

**Keywords:** atrial fibrillation, cost of illness, cost trajectories, hierarchical clustering, patient cluster

## Abstract

**Aims:** Evidence on long-term costs of atrial fibrillation (AF) and associated factors is scarce. As part of the Swiss-AF prospective cohort study we aimed to characterise AF costs and their development over time, and to assess specific patient clusters and their cost trajectories.

**Methods:** Swiss-AF enrolled 2,415 patients with variable duration of AF between 2014 and 2017. Patient clusters were identified using hierarchical cluster analysis of baseline characteristics. Ongoing yearly follow-ups include health insurance clinical and claims data. An algorithm was developed to adjudicate costs to AF and related complications.

**Results:** Hierarchical analysis identified three patient clusters. “Cardiovascular-dominated” (CV-dominated) patients had the highest proportions of prior myocardial infarction and presence of diabetes. “Heart failure-dominated” (HF-dominated) patients had the highest occurrence of heart failure and permanent AF. “Isolated symptomatic” (IS) patients were younger and had the highest occurrence of paroxysmal AF. A subpopulation of 1,024 Swiss-AF patients with available claims data was followed up for a median [interquartile range] of 3.24 [1.09] years. Average yearly AF-adjudicated costs amounted to CHF 5,679, remaining stable across the observation period. CV-dominated (N = 253 with claims data) and HF-dominated patients (N = 185) depicted similarly high costs across all cost outcomes, the IS (N = 586) patients accrued the lowest costs.

**Conclusion:** Our results highlight three well-differentiated patient clusters with specific costs that could be used for stratification in both clinical and economic studies. Patient characteristics associated with adjudicated costs as well as cost trajectories may enable an early understanding of the magnitude of upcoming AF-related healthcare costs.

**What is already known on this topic:** Atrial fibrillation (AF) is a complex disease and constitutes a major economic and societal challenge due to its high prevalence worldwide.

**What this study adds:** This study, based on a large prospective cohort study, provides evidence on real-world AF costs and their development over time. Data-derived patient clusters are linked to costs and their respective cost trajectories are assessed.

**How this study might affect research, practice or policy:** The identified patient clusters and their characteristics may help clinicians and payers to gain an early insight and understanding of the magnitude of the expected AF-related healthcare costs.

## Introduction

Atrial fibrillation (AF) is the most prevalent arrhythmia worldwide,^1^ and its prevalence is expected to double by 2050.^2,3^ AF patients face increased risk of stroke, cognitive dysfunction, and impaired quality of life.^4,5^

AF also constitutes a major economic and societal challenge. However, only a limited number of AF cost-of-illness studies^6^ is available, indicating that costs are predominantly driven by AF-related hospitalisations.^7^ Hospitalisation costs were reported to amount to 50-70% of total costs,^8^ while medication costs were comparatively low.^9^ Drivers of hospitalisations in AF patients were identified to be multifaceted and diverse.^10,11^ They include age, cardiovascular diseases like previous history of heart failure and myocardial infarction, non-cardiovascular conditions, i.e. sleep apnoea and chronic kidney disease, as well as lifestyle factors.

Despite these insights, evidence on real-world AF-related costs and their components remains scarce. Most results stem from retrospective studies. Even less evidence is available on cost trajectories over time, especially in prevalent AF patients. Patient characterisation based on AF subtypes is known to not encompass the full heterogeneity of AF,^12,13^ and data-driven characterisation of patients has not yet been used for costing.

The Swiss-AF Economics project is part of the prospective Swiss Atrial Fibrillation cohort study^14,15^ (Swiss-AF) and aims to assess AF-related economic burden. Clinical data are combined with health insurance claims data to characterise the Swiss-AF study population from an economic perspective. Focusing on the direct medical cost per patient from the perspective of the Swiss statutory health insurance system, the present work aims to: i) describe AF costs and their trajectories, and ii) to build data-driven patient clusters and assess their respective cost trajectories.

## Methods

### Study Design and Data Sources

Swiss-AF is an ongoing prospective multicentre observational cohort study across 13 clinical centres in Switzerland. The detailed study setup and methodology have been described earlier.^14^ In short, 2,415 patients were enrolled between April 2014 and August 2017; this analysis used a 2014-2020 data cut. Eligibility criteria for Swiss-AF were based on a history of documented AF (paroxysmal, persistent, or permanent) and being older than 65 years; 228 patients were enrolled at age 45-64 years to enhance the study of socio-economic aspects of AF.

Health economic data include medical resource use at the enrolling centres and health insurance claims data from four cooperating insurance companies, covering 1,024 patients (42.4% of the study population). The claims data cover all services submitted for reimbursement by the Swiss statutory health insurance. The statutory health insurance is obligatory for all residents and has a comprehensive benefit package covering inpatient and outpatient services. An insured person can choose from a variety of deductibles (CHF 300–2,500), where a higher deductible leads to a lower premium.

Claims relating to inpatient services were based on the Swiss flat-fee reimbursement scheme for acute care hospitalisations (SwissDRG^16^). Data related to inpatient services in rehabilitation clinics were based on tariffs per day negotiated between each service provider and insurer, and claims related to nursing home care services were based on daily contributions fixed by the Swiss Federal Council. Outpatient services were covered with detailed information for each cost component. Relevant comorbidities not collected by the Swiss-AF study were derived from the drug utilisation present in the claims data, using the pharmaceutical cost groups (PCGs) method.^17^

### Outcome Measures

Our main outcome of interest was total AF-adjudicated costs from the Swiss statutory health insurance perspective. Costs in AF patients are typically partially related to AF and its potential complications, partially related to other comorbidities. To distinguish these components, an adjudication algorithm was developed through an iterative process involving several clinical experts of the Swiss-AF study centres. The adjudication algorithm (https://gitlab.uzh.ch/helena.aebersold/patient-clusters-and-cost-trajectories.git) combined clinical events, e.g. date of a bleeding event, with information from the health insurance claims, e.g. SwissDRG based. Inpatient costs were either non-related or adjudicated to one of five groups: (1) AF, (2) stroke or transient ischemic attack (TIA), (3) bleeding, (4) fall, (5) heart failure. Similarly, outpatient costs were either non-related or classified into one of six groups: (1) AF without medication, (2) AF medication, (3) stroke or TIA without medication, (4) stroke or TIA medication, (5) bleeding, (6) fall. As it was not possible to distinguish between complications (e.g. strokes) caused by AF or other reasons, complication costs were considered in full.

We thus considered the sum of all costs adjudicated to groups (1) to (6) as total AF-adjudicated costs. Secondary outcomes included AF-adjudicated inpatient costs, AF-adjudicated outpatient costs excluding drug costs, and AF-adjudicated outpatient drug costs. We also considered total costs, i.e., the sum of total AF-adjudicated costs and total non-adjudicated costs, and equivalent sub-categories. All outcome variables and their relation can be found in **Figure S1** in the supplement.

### Covariates

Baseline covariates, measured at enrolment, included the following types:

1. patient characteristics: age, sex, body mass index (BMI), AF type, AF symptoms, time since AF diagnosis, risk of stroke for non-valvular atrial fibrillation (CHA_²_DS_²_-VASc score).
2. medical history: previous major bleeding, stroke or TIA, systemic embolism, heart failure, myocardial infarction, diabetes, hypertension, renal insufficiency, sleep apnoea.
3. treatments and implanted devices: percutaneous transluminal coronary angioplasty (PTCA), coronary artery bypass grafting (CABG), electro-conversion, pulmonary vein isolation (PVI), loop recorder, pacemaker (PM), cardiac resynchronisation therapy (CRT) with or without ICD, implantable cardioverter defibrillator (ICD).
4. anticoagulation medication: vitamin K antagonist (VKA), direct-acting oral anticoagulant (DOAC).
5. other medication: antiplatelets, aspirin, statins, diuretics, beta-blockers, digoxin.
6. socioeconomic factors: education, greater area of residence, smoking, alcohol consumption.

### Statistical analysis

First, the characteristics of the included Swiss-AF patients were described. Baseline characteristics were presented with mean and standard deviation for normally distributed variables, median and interquartile range for continuous non-normally distributed variables, and N (%) for categorical ones. Kaplan-Meier curves of overall survival were used to depict mortality and cohort attrition.

Second, the costs and their trajectories since enrolment were described. Cost components are shown as bar charts and boxplot distributions of mean annual costs. Cost trajectories are depicted as line plots with mean estimates and 95% confidence intervals. All costs considered individual follow-up times and were aggregated to a yearly level. Given the relative stability of prices over the observation period, costs were taken as reported. Costs as a function of time since diagnosis were also explored graphically. Third, to gain an understanding of different patterns in the Swiss-AF population, clusters of patients and their respective cost composition and cost trajectories were determined. A hierarchical cluster analysis was run. The unsupervised learning model^18^ considered the full Swiss-AF population and all baseline covariates except greater area of residence. Gower distances were used with a complete linkage method, choosing optimal clusters according to elbow and silhouette methods.^18^ Differences in baseline characteristics between patient clusters were compared with standardised mean difference, t-test for continuous and roughly normally distributed variables, Mann-Whitney U test for continuous non-normally distributed variables, and fisher’s exact test for categorical variables. Cluster-specific overall survival was compared using Kaplan-Meier curves and age-adjusted cox regression. A complementary multivariable regression-based cost analysis is reported in the supplement (**Figures S7-S8, Tables S5-S8**).

A 5% significance level was pre-specified, and no multiple hypothesis testing adjustments were performed due to the exploratory nature of the study. Given the usage of baseline data and observed claims there were no missing data. The cost analyses conditioned on patients being alive. Data management, variable derivations and all statistical analyses were conducted using R version 3.6.3.

## Results

### Patient population

Out of 2,415 Swiss-AF patients, 1,024 (42.4%) had available claims data and were included in the cost analyses. Baseline characteristics are shown in Fehler! Verweisquelle konnte nicht gefunden werden.. The mean age at baseline was 73 ± standard deviation (SD) 8 years and 28% of the participants were female. On average, patients were diagnosed with AF 6.3 years prior to enrolment, and the majority had AF symptoms (62%). Patients without available claims data, additionally used in the hierarchical cluster analysis, had comparable characteristics (Fehler! Verweisquelle konnte nicht gefunden werden. in the supplement). **Figure S2** in the supplement depicts the average monthly cost evolution alongside the Kaplan-Meier curve of overall survival, across the full observation period of 2014 to 2020. For subsequent analyses, individual patient follow-up was censored at five years after enrolment (median follow-up: 3.24 years), given sparse data thereafter.

### Cost structures and cost trajectories

Mean annual costs and cost trajectories for different cost outcomes are shown in **Figure 1**. More details on all cost outcomes and costs by time since AF diagnosis can be found in the supplement (**Tables S2, Figures S3-S4**). The average total per-patient costs amounted to CHF 19,037 yearly, of which roughly half corresponded to outpatient services. Inpatient costs appeared to be higher than outpatient costs only during the first year, remaining stable and at a similar level thereafter.

**Figure 1.**
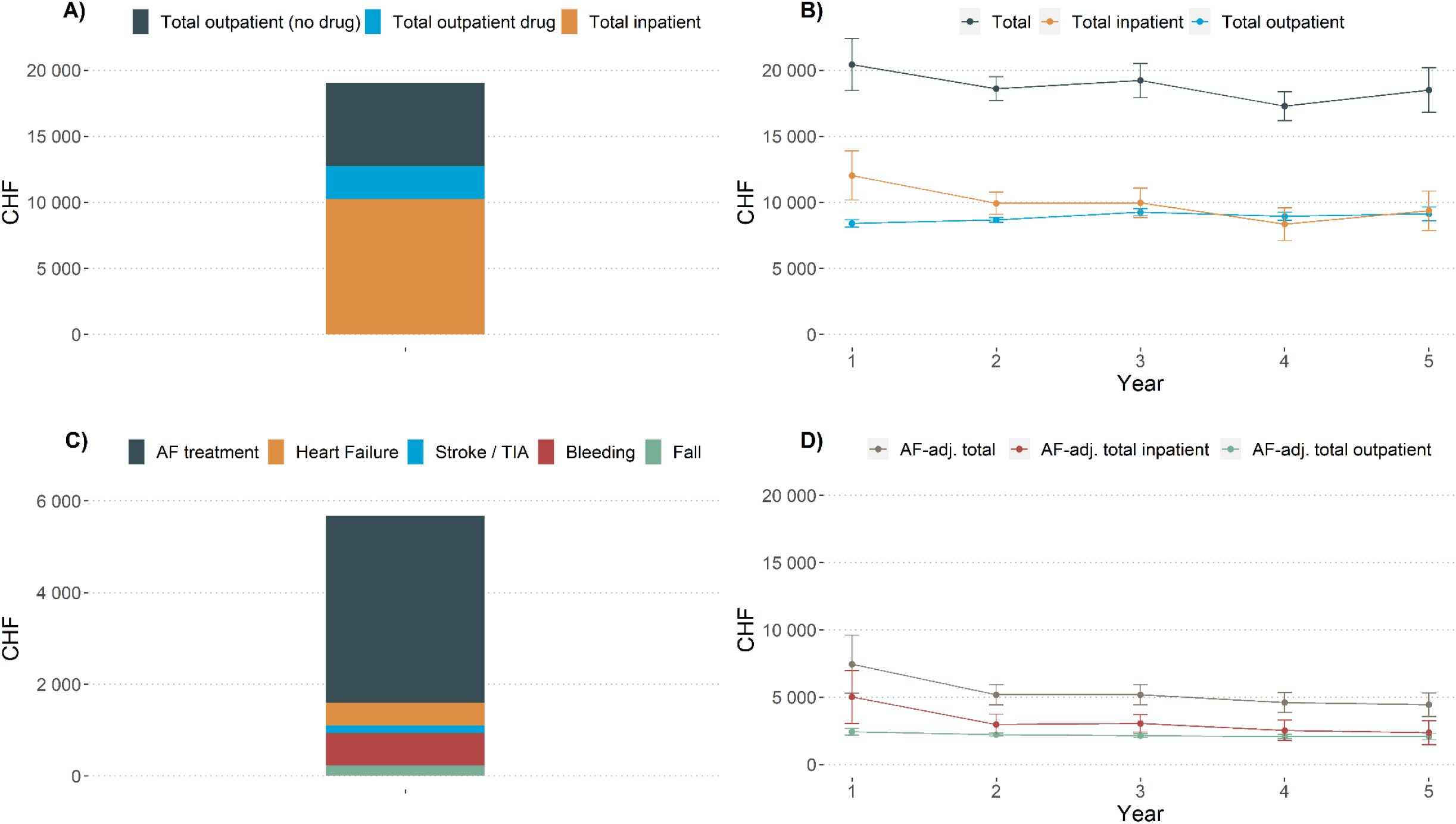
Cost compositions and cost trajectory of different outcome variables. A) Composition of mean annual total costs. B) Cost trajectory of total, total inpatient and total outpatient costs during five years of follow-up. C) Composition of mean annual AF-adjudicated total costs. AF treatment costs include AF medication, and stroke / TIA costs include stroke / TIA medication. All cost components include inpatient and outpatient services, except for heart failure. Heart failure costs were calculated only from inpatient services. D) Cost trajectory of AF-adjudicated total, AF-adjudicated total inpatient and AF-adjudicated total outpatient costs during five years of follow-up. *Notes:* Abbreviations: adj.: adjudicated, AF: atrial fibrillation.

Total AF-adjudicated costs amounted to CHF 5,679 annually, roughly 30% of the total costs. AF-adjudicated costs consisted mainly of AF treatment costs (CHF 4,078), followed by costs of bleeding (CHF 696) and heart failure (CHF 494). As the total costs, the AF-adjudicated costs remained stable across the whole analysed period of five years. AF-adjudicated inpatient costs were higher than AF-adjudicated outpatient costs in the first year.

### Patient clusters and their cost trajectories

A total of three hierarchical analysis-based clusters were identified to be optimal according to the elbow and silhouette methods. Baseline characteristics by cluster are shown in **Table 1**. Cluster-specific Kaplan-Meier curves of overall survival and results of age-adjusted cox regression are depicted in **Figure S5** and **Table S3**.

**Table 1.**
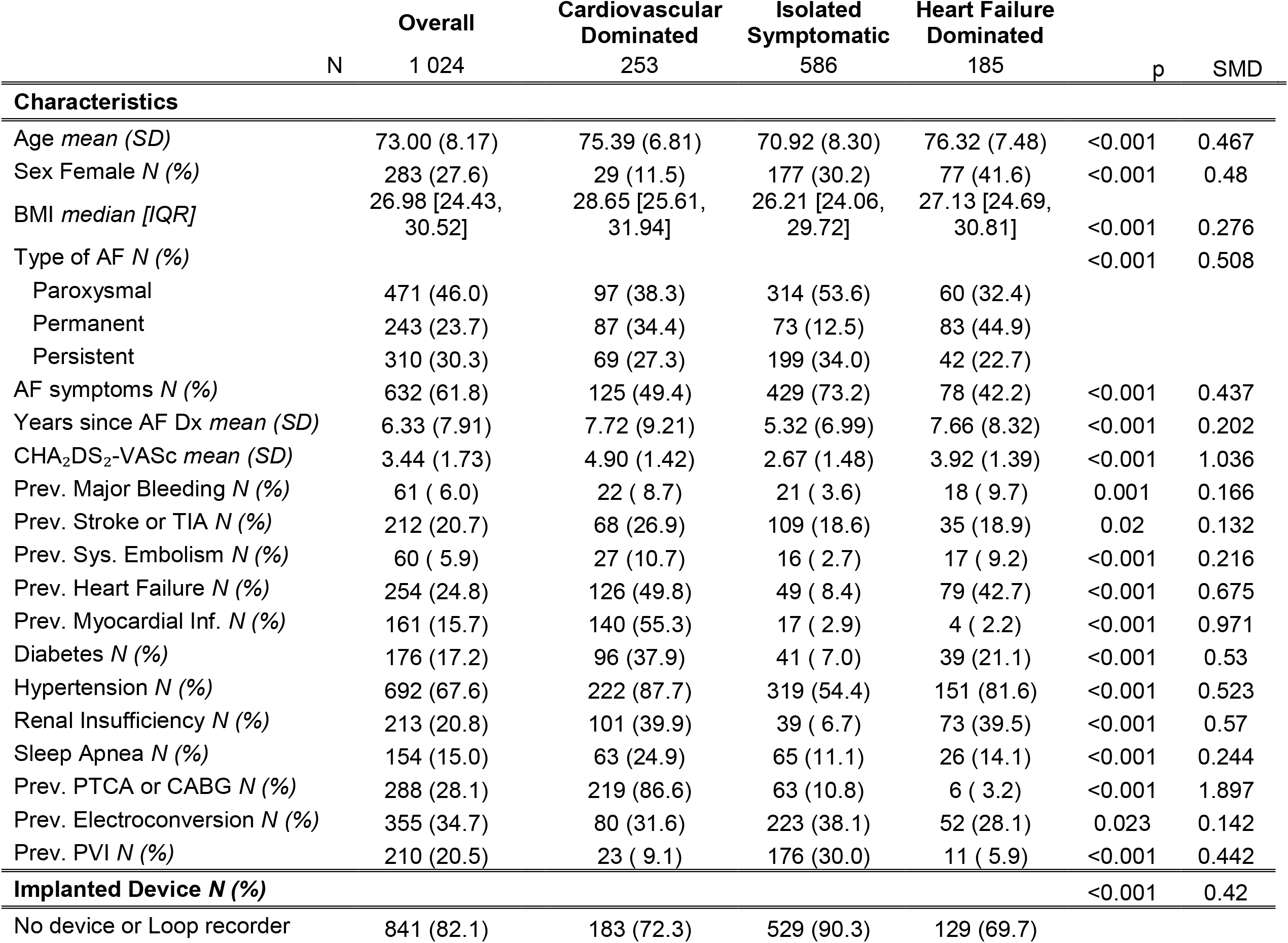
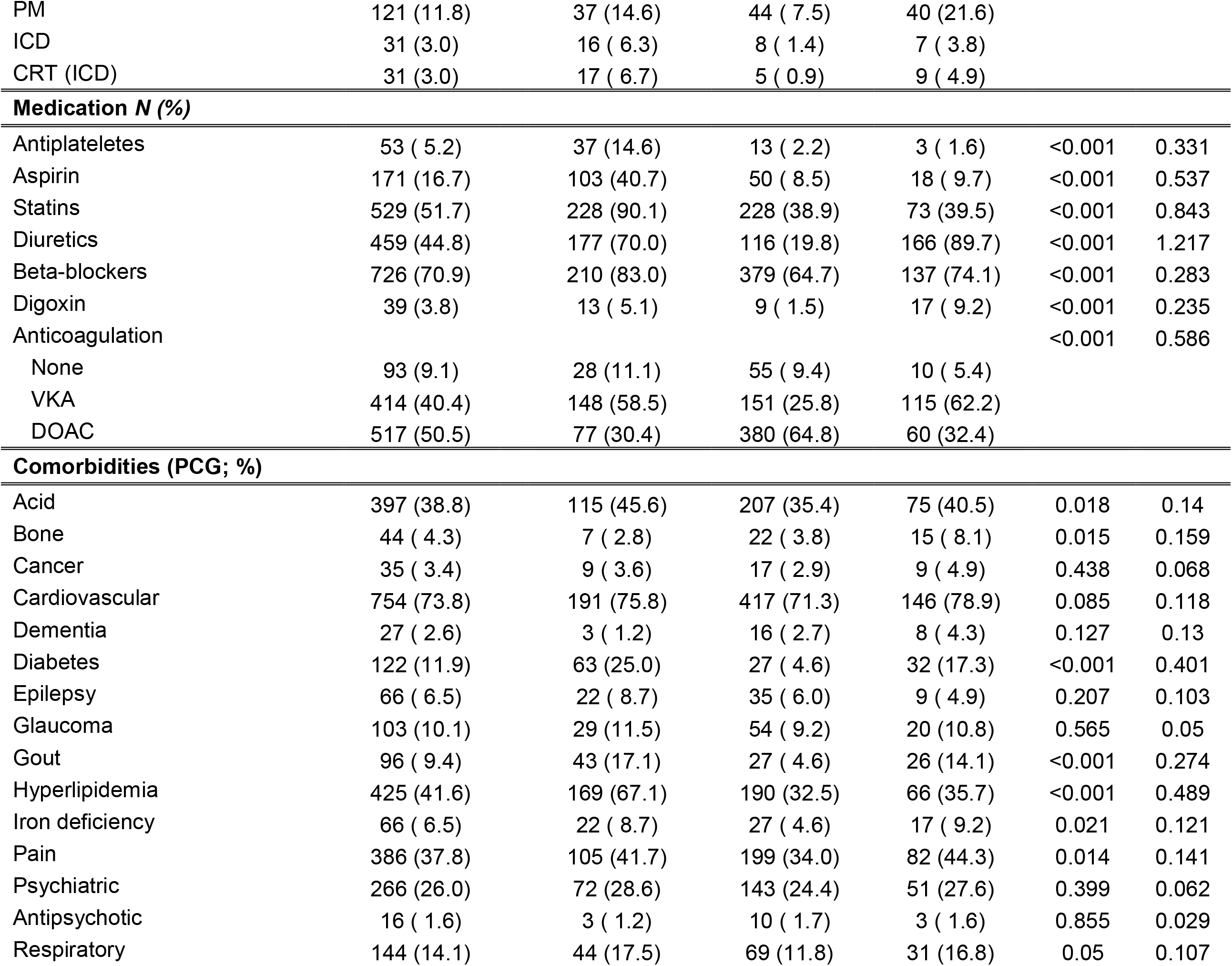
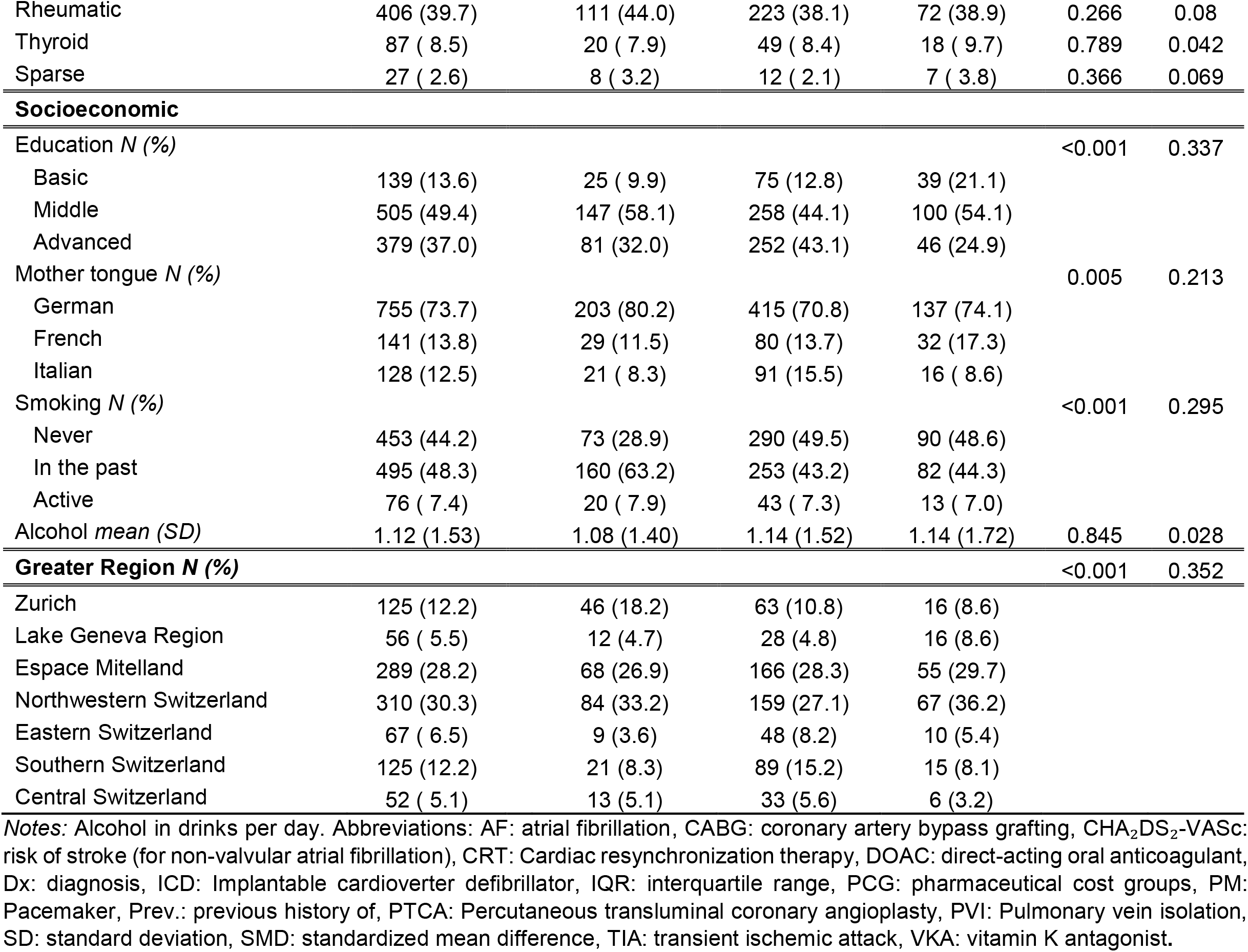
Baseline characteristics of all included AF patients and each cluster.

The first cluster included 253 patients and may be characterised as “cardiovascular dominated” (CV-dominated). Most patients in this cluster had previous PTCA or CABG (87%), and we observed the highest proportions of prior myocardial infarction (55%), heart failure (50%), and stroke or TIA (27%). There was also a high presence of diabetes (38%), hyperlipidaemia (67%), and past smoking (63%). Until the five-year follow-up 55 (21.7%) were deceased and 11 (4.3%) lost.

The second cluster included 586 patients we characterise as being “isolated symptomatic” (IS). Patients in this cluster were the youngest among the three clusters (mean age: 71). They had the highest occurrence of paroxysmal AF (54%) and presence of AF symptoms (73%). However, they had fewer medical history items present, fewer comorbidities, low medication use, and had been diagnosed with AF more recently (mean years: 5.3). Until the 5-year follow-up 36 (6.1%) were deceased and 14 (2.4%) lost.

The third cluster included 185 “heart failure dominated” (HF-dominated) patients. These patients were characterised by the highest occurrence of permanent AF (45%), a high presence of heart failure (43%), and the highest proportion with a pacemaker (22%). Moreover, they featured the lowest presence of AF symptoms (42%), had practically no previous myocardial infarctions (2.2%), low prevalence of diabetes (21%), and low aspirin usage (9.7%). Many used diuretics (90%). Until the 5-year follow-up 35 (18.9%) were deceased and 12 (6.5%) lost.

Cost trajectories for each cluster are shown in **Figure 2**. More details on the cost outcomes by cluster can be found in **Table S4** and **Figure S6**. Throughout all cost trajectories, the IS patients accrued the lowest costs. Moreover, the pattern in the IS group resembled the overall cost trajectory (see **Figure 1**): after an initial high, all costs remained stable across the observation period. Compared to the HF-dominated cluster, the CV-dominated patients depicted slightly higher AF-adjudicated costs. The results are consistent with the results of the complementary multivariable regression-based cost analysis, which provided estimates of average covariate effects across all patients (**Figures S7-S8, Tables S5-S8**).

**Figure 2.**
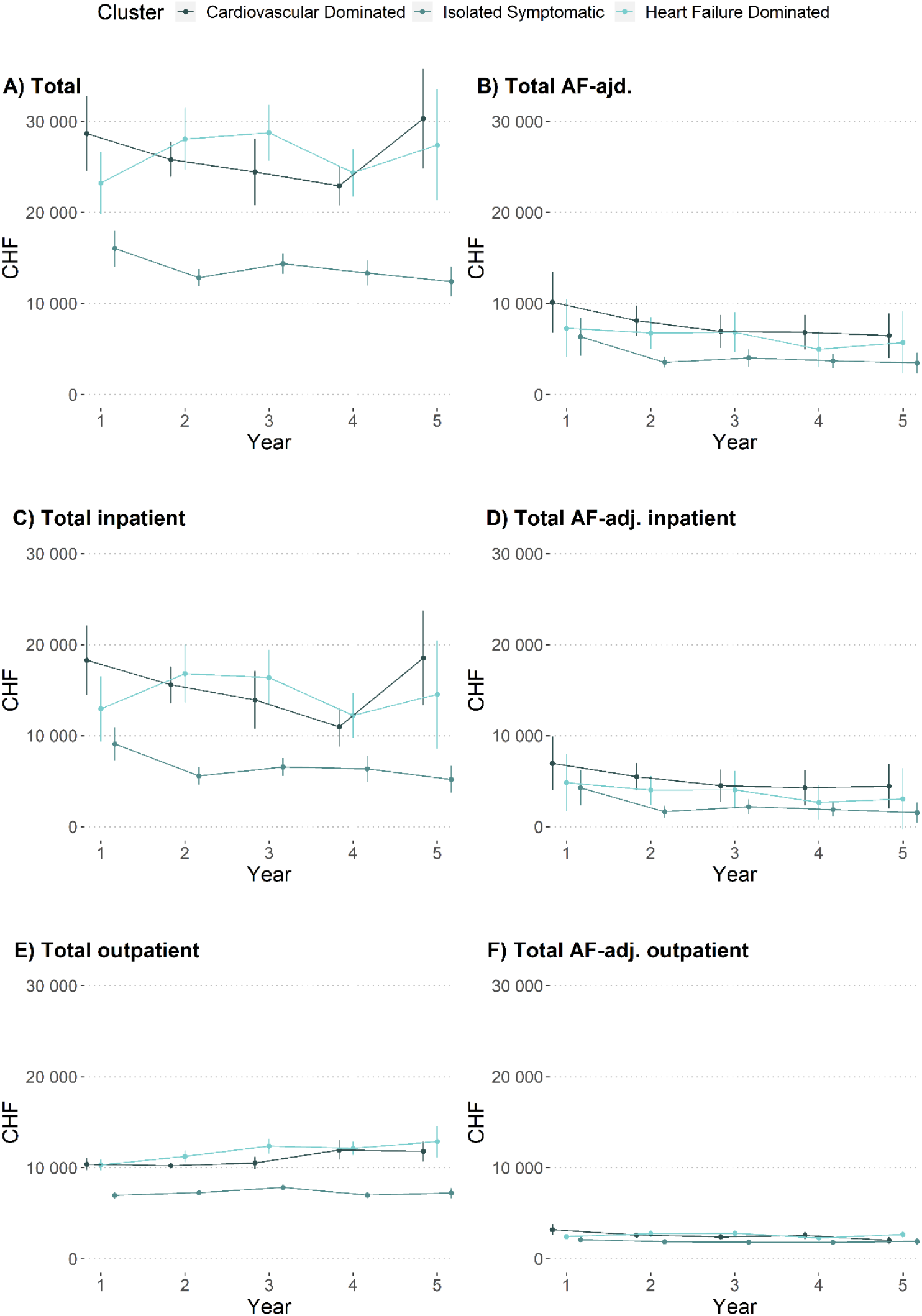
Cost trajectories by patient clusters. Three patient clusters were identified: Cardiovascular Dominated, Isolated Symptomatic AF, and Heart Failure Dominated. The cost trajectory for different cost outcomes are shown for a follow-up period of five years: A) total costs, B) total AF-adjudicated costs, C) total inpatient costs, D) total AF-adjudicated inpatient costs, E) total outpatient costs, and F) total AF-adjudicated outpatient costs. *Notes:* Abbreviations: adj.: adjudicated, AF: atrial fibrillation.

## Discussion

This study presents recent evidence on real-world AF costs. To the best of our knowledge, it is the first to link data-derived patient clusters with cost trajectories. We observed mean annual and AF-adjudicated healthcare costs to remain stable over several years. Furthermore, among three clinically well-differentiated patient clusters cost levels differed but were also stable throughout the observation period.

Mean annual total costs amounted to CHF 19,037 per AF patient (∼23% of the Swiss Gross Domestic Product per capita), whereof CHF 5,679 were attributable to AF and its complications. Across the entire observation period, these costs remained relatively stable. Inpatient and outpatient costs were of similar magnitude; this applied to both, AF-adjudicated and non-adjudicated costs. This finding is in line with existing literature, where inpatient costs incurred at least half of the total costs.^19–22^ The observation of a stable proportion across five years, also apparent in AF-adjudicated costs, has not been previously described.

Of the AF-adjudicated costs, 71.8% were directly related to AF, followed by bleeding, heart failure, fall, and stroke / TIA costs. In the literature so far, AF-related costs were mostly assessed through various excess cost calculations^23^ and a clinically-based bottom-up approach was missing. Even though our adjudication algorithm was unable to distinguish between the costs of care episodes triggered by AF versus by other causes, we believe that this bottom-up approach contributes to a more realistic understanding of AF and related costs. Also, in contrast to most studies,^19,21,24,25^ our observed contribution of AF-adjudicated costs to overall healthcare costs was smaller. One reason may be that we analysed prevalent AF patients while the literature focussed mainly on incident patients.

Three well-differentiated patient clusters resulted from our analysis, with distinct cost trajectories: CV-dominated with rather high costs, HF-dominated with similarly high costs, and IS with substantially lower costs. Some attempts to cluster the heterogeneity of AF patients were reported previously, but not linked with cost trajectories.^12,26–28^ To facilitate an understanding of expected patient trajectories, we based our analysis on baseline characteristics.

Our first cluster, the CV-dominated patients, featured many strong risk factors for cardiovascular diseases (e.g., high rates of past smokers, low rate of never smokers, high prevalence of diabetes) and high cardiovascular drug use (i.e. aspirin, statin, and beta-blocker). The high rates of cardiovascular revascularisation procedures, myocardial infarction and stroke/TIA suggested underlying cardiovascular disease. These characteristics are roughly comparable with those of the Atherosclerotic Comorbid AF cluster found in two other cluster studies.^12,26^ The CV-dominated patients accrued substantial costs, especially AF-adjudicated costs, which remained stable at a high level across time. This result is strongly consistent with the complementary multivariate approach.

The second cluster, representing IS, showed the lowest costs among the three clusters. Patients were noticeably younger, had fewer comorbidities, but high rates of previous cardioversion, AF symptoms, and paroxysmal AF. This cluster has partially similar characteristics as the Younger Paroxysmal AF cluster identified in a Japanese cohort^29^ and the Low Comorbidity cluster identified in a US cohort^12^. Differences in cultural and clinical settings, and in available descriptors of patient characteristics may have limited the degree of similarity. Considering the results of the multivariable regression-based analysis, this cluster seems to possess very few covariates that drive the costs significantly above a base level, explaining the low cost level.

The third cluster, HF-dominated, featured similarly high costs as the CV-dominated, as well as similarly high rates of heart failure. However, patients had almost no prior myocardial infarction, little diabetes, low aspirin intake, yet high diuretics intake. This may be consistent with heart failure caused by underlying causes other than cardiovascular disease. The patients also featured a relatively low prevalence of covariates associated with higher AF-adjudicated costs but were at the same cost level as the CV-dominated cluster. A possible explanation for this result may be unplanned hospitalisations and complications due to the ongoing challenge to treat patients with heart failure. This specific cluster has not yet been reported by other AF cluster studies.^12,26^

The prospective cohort study patients we report on differ from the patient populations underlying recent AF costs studies in four ways, which has implication for the interpretation of the results. Firstly, most studies to-date relied on administrative databases^21,25,30^; secondly, they were often retrospective^19,21,25,27^; and thirdly, had short follow-up times between 0.5 to 3 years^19,24,29^. In combining clinical and health insurance claims data prospectively, the present analysis adds detailed insight into the longer-term healthcare costs of AF patients. Fourthly, the present study examined prevalent patients with varying disease duration (including 13% enrolled within six months after first AF diagnosis). Even though the Swiss-AF study population cannot be regarded as a representative sample of AF patients, it provides insights on the longer-term cost impact of AF in a substantially wider spectrum of AF patients than previous studies.

There are several limitations inherent to our study that require discussion. First, the data structure is highly complex and patients in Swiss-AF are very heterogeneous. We tried to account for the heterogeneity with the clustering method, and covariate selection was based on the current literature as well as discussions with clinicians from the Swiss-AF study centres. Second, as the study population is not truly representative of all Swiss AF patients, a typical ‘average’ AF cost could not be reliably deducted. Further work will approach this topic with a general population-based, matched control population. Third, given deductible levels in the Swiss statutory health insurance system, the possibility that not all claims were handed in for reimbursement exists, which may have led to an underestimation of true costs. However, as AF patients have substantial costs, it is rather unlikely that claims were not submitted. A fourth limitation lies in the adjudication algorithm, which was developed carefully and considered clinical information together with claims data. Still, it may not fully reflect the true causation of costs by AF and its complications, and could not distinguish which specific complications, e.g. strokes, were truly due to AF or not.

In conclusion, this study analysed real-world AF costs, patient clusters and their cost trajectories. We observed mean annual AF-adjudicated costs of roughly CHF 6,000 per patient and stable relative contributions of different cost categories during five years. Three well-differentiated patient clusters were identified using a hierarchical clustering method: CV-dominated, HF-dominated and IS patients. The cost levels for AF-adjudicated and overall healthcare costs depended on patient cluster but remained again stable over time. CV-dominated patients accrued the highest costs and HF-dominated patients only slightly lower ones, while IS patients had substantially lower costs. These insights may help clinicians and payers to develop an early understanding of the magnitude of to be expected AF-related healthcare costs.

## Data Availability

The patient informed consent forms state that the data, containing personal and medical information, are exclusively available for research institutions in an anonymized form and are not allowed to be made publicly available. Researchers interested in obtaining the data for research purposes can contact the Swiss-AF scientific lead. Contact information is provided on the Swiss-AF website (http://www.swissaf.ch/contact.htm). Authorization of the responsible ethics committee is mandatory before the requested data can be transferred to external research institutions.

## Acknowledgements

We thank Lukas Kauer from the health insurance CSS, Pascal Godet from the health insurance Sanitas, Beat Brüngger and Andri Signorell from the health insurance Helsana, and the health insurance KPT for providing the claims data for this study. The full list of Swiss-AF investigators is provided in the supplementary material.

## Ethics and Data availability statement

The Swiss-AF study protocol was approved by the local ethics committee (Ethikkommission Nordwest-und Zentralschweiz), and written informed consent was obtained from each participant. The patient informed consent forms state that the data, containing personal and medical information, are exclusively available for research institutions in an anonymized form and are not allowed to be made publicly available. Researchers interested in obtaining the data for research purposes can contact the Swiss-AF scientific lead. Contact information is provided on the Swiss-AF website (http://www.swissaf.ch/contact.htm). Authorization of the responsible ethics committee is mandatory before the requested data can be transferred to external research institutions.

## References

1. Patel, N. J., Atti, V., Mitrani, R. D., Viles-Gonzalez, J. F. & Goldberger, J. J. Global rising trends of atrial fibrillation: a major public health concern. Heart 104, 1989–1990 (2018).

2. Krijthe, B. P. et al. Projections on the number of individuals with atrial fibrillation in the European Union, from 2000 to 2060. European Heart Journal 34, 2746–2751 (2013).

3. Miyasaka, Y. et al. Secular Trends in Incidence of Atrial Fibrillation in Olmsted County, Minnesota, 1980 to 2000, and Implications on the Projections for Future Prevalence. Circulation 114, 119–125 (2006).

4. Conen, D. et al. Risk of Death and Cardiovascular Events in Initially Healthy Women With New-Onset Atrial Fibrillation. JAMA 305, 2080–2087 (2011).

5. Stewart, S., Murphy, N. F., Walker, A., McGuire, A. & McMurray, JJ. Cost of an emerging epidemic: an economic analysis of atrial fibrillation in the UK. Heart 90, 286–292 (2004).

6. Wodchis, W. P., Bhatia, R. S., Leblanc, K., Meshkat, N. & Morra, D. A Review of the Cost of Atrial Fibrillation. Value in Health 15, 240–248 (2012).

7. Ball, J., Carrington, M. J., McMurray, J. J. V. & Stewart, S. Atrial fibrillation: Profile and burden of an evolving epidemic in the 21st century. International Journal of Cardiology 167, 1807–1824 (2013).

8. Wolowacz, S. E., Samuel, M., Brennan, V. K., Jasso-Mosqueda, J. G. & van Gelder, I. C. The cost of illness of atrial fibrillation: a systematic review of the recent literature. Europace 13, 1375–1385 (2011).

9. Wong, C. X., Brooks, A. G., Leong, D. P., Roberts-Thomson, K. C. & Sanders, P. The Increasing Burden of Atrial Fibrillation Compared With Heart Failure and Myocardial Infarction: A 15-Year Study of All Hospitalizations in Australia. Arch Internal Medicine 172, 739–741 (2012).

10. Bhat, A. et al. Drivers of hospitalization in atrial fibrillation: A contemporary review. Heart Rhythm 17, 1991–1999 (2020).

11. Steinberg, B. A. et al. Drivers of hospitalization for patients with atrial fibrillation: Results from the Outcomes Registry for Better Informed Treatment of Atrial Fibrillation (ORBIT-AF). American Heart Journal 167, 735-742.e2 (2014).

12. Inohara, T. et al. Association of of Atrial Fibrillation Clinical Phenotypes With Treatment Patterns and Outcomes: A Multicenter Registry Study. JAMA Cardiology 3, 54–63 (2018).

13. Lip, G., Nieuwlaat, R., Pisters, R., Lane, D. & Crijns, H. Refining clinical risk stratification for predicting stroke and thromboembolism in atrial fibrillation using a novel risk factor-based approach: the euro heart survey on atrial fibrillation. Chest 137, 263–272 (2010).

14. Conen, D. et al. Design of the Swiss Atrial Fibrillation Cohort Study (Swiss-AF): structural brain damage and cognitive decline among patients with atrial fibrillation. Swiss Medical Weekly 147, (2017).

15. Conen, D. et al. Relationships of Overt and Silent Brain Lesions With Cognitive Function in Patients With Atrial Fibrillation. J Am Coll Cardiol 73, 989–999 (2019).

16. SwissDRG. https://www.swissdrg.org/de/akutsomatik/swissdrg-system-1002021 (2021).

17. Huber, C. A., Szucs, T. D., Rapold, R. & Reich, O. Identifying patients with chronic conditions using pharmacy data in Switzerland: an updated mapping approach to the classification of medications. BMC Public Health 13, 1–10 (2013).

18. Hastie, T., Tibshirani, R. & Friedman, J. Springer Series in Statistics The Elements of Statistical Learning Data Mining, Inference, and Prediction.

19. Reinhold, T., Lindig, C., Willich, S. N. & Brüggenjürgen, B. The costs of atrial fibrillation in patients with cardiovascular comorbidities--a longitudinal analysis of German health insurance data. Europace 13, 1275–1280 (2011).

20. Zoni-Berisso, M. et al. The cost of atrial fibrillation in Italy: a five-year analysis of healthcare expenditure in the general population. From the Italian Survey of Atrial Fibrillation Management (ISAF) study. European Review for Medical and Pharmacological Sciences 21, 175–183 (2017).

21. Kim, M., Lin, J., Hussein, M., Kreilick, C. & Battleman, D. Cost of atrial fibrillation in United States managed care organizations. Advances in Therapy 26, 847–857 (2009).

22. Ringborg, A. et al. Costs of atrial fibrillation in five European countries: Results from the Euro Heart Survey on atrial fibrillation. Europace 10, 403–411 (2008).

23. Becker, C. Cost-of-illness studies of atrial fibrillation: methodological considerations. Expert Review of Pharmacoeconomics & Outcomes Research 14, 661–684 (2014).

24. Johnsen, S. P., Dalby, L. W., Täckström, T., Olsen, J. & Fraschke, A. Cost of illness of atrial fibrillation: a nationwide study of societal impact. BMC Health Services Research 17, 1–8 (2017).

25. Kim, M., Johnston, S., Chu, B., Dalal, M. R. & Schulman, K. Estimation of total incremental health care costs in patients with atrial fibrillation in the United States. Circulation. Cardiovascular Quality and Outcomes 4, 313–320 (2011).

26. Inohara, T. et al. A Cluster Analysis of the Japanese Multicenter Outpatient Registry of Patients With Atrial Fibrillation. The American Journal of Cardiology 124, 871–878 (2019).

27. Suzuki, S. et al. Identifying risk patterns in older adults with atrial fibrillation by hierarchical cluster analysis: A retrospective approach based on the risk probability for clinical events. IJC Heart & Vasculature 37, 100883 (2021).

28. Karwath, A. et al. Redefining β-blocker response in heart failure patients with sinus rhythm and atrial fibrillation: a machine learning cluster analysis. The Lancet 398, 1427–1435 (2021).

29. Boccuzzi, S. et al. Retrospective study of total healthcare costs associated with chronic nonvalvular atrial fibrillation and the occurrence of a first transient ischemic attack, stroke or major bleed. Curr Med Res Opin 25, 2853–2864 (2009).

30. Bennell, M. C. et al. Identifying predictors of cumulative healthcare costs in incident atrial fibrillation: A population-based study. J Am Heart Assoc 4, (2015).

