## supplement for "Patient Clusters and Cost Trajectories in Atrial Fibrillation: Evidence from the Swiss Atrial Fibrillation Cohort"

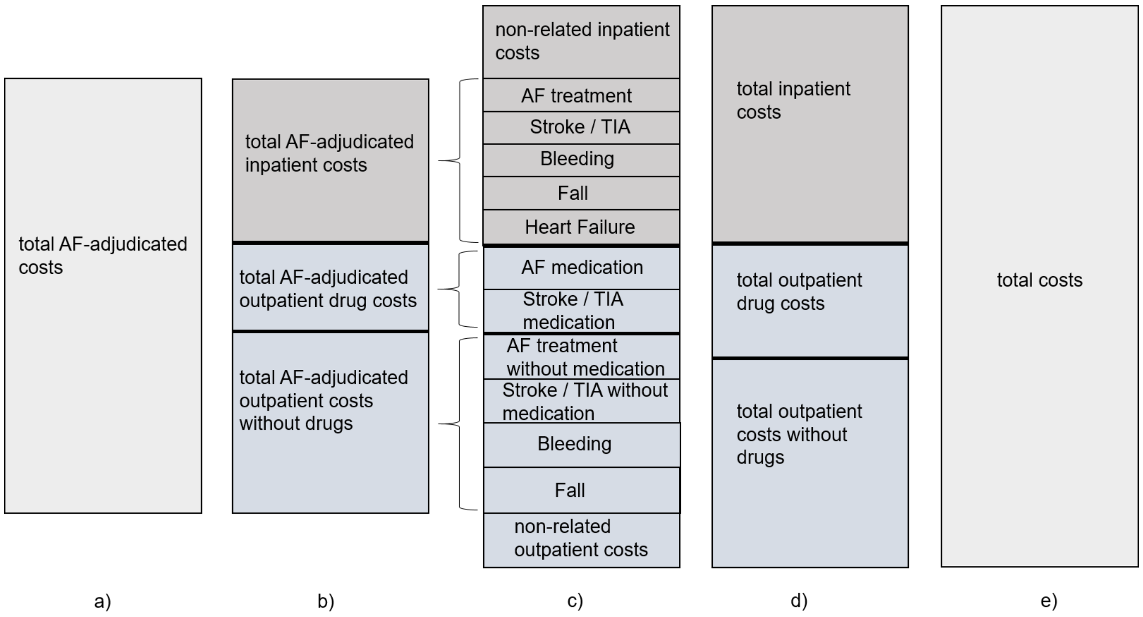

**Figure S1. Outcome definition scheme.** Main outcome of interest was (a) total AF-adjudicated costs, which is (b) the sum of total AF-adjudicated inpatient costs and total AF-adjudicated outpatient costs. Total inpatient costs (d) are either adjudicated into five groups or non-related (c), while total outpatient costs (d) are either adjudicated into six groups or non-related (c). The sum of all costs account for total costs (e).

**Table S1. Baseline characteristics of patients with and without claims data.**

| | Claims | No claims | | |
|---|---|---|---|---|
| n | 1 024 | 1 391 | p | SMD |
| **Characteristics** | | | | |
| Age *mean (SD)* | 73.00 (8.17) | 73.42 (8.61) | 0.232 | 0.049 |
| Sex Female *N (%)* | 283 (27.6) | 379 (27.2) | 0.868 | 0.009 |
| BMI *median [IQR]* | 26.98 [24.43, 30.52] | 26.99 [24.34, 30.25] | 0.849 | 0.001 |
| Type of AF *N (%)* | | | 0.536 | 0.046 |
| Paroxysmal | 471 (46.0) | 611 (43.9) | | |
| Permanent | 243 (23.7) | 353 (25.4) | | |
| Persistent | 310 (30.3) | 427 (30.7) | | |
| AF symptoms *N(%)* | 632 (61.8) | 862 (62.0) | 0.964 | 0.004 |
| Years since AF Dx *mean (SD)* | 6.33 (7.91) | 6.06 (7.49) | 0.399 | 0.035 |
| $CHA_2DS_2$-VASc score *mean (SD)* | 3.44 (1.73) | 3.50 (1.68) | 0.411 | 0.034 |
| Prev. Major Bleeding *N (%)* | 91 ( 6.5) | 61 (6.0) | 0.617 | 0.024 |
| Previous Stroke or TIA *N (%)* | 212 (20.7) | 267 (19.2) | 0.396 | 0.037 |
| Prev. Sys. Embolism *N (%)* | 67 ( 4.8) | 60 (5.9) | 0.299 | 0.046 |
| Prev. Heart Failure *N (%)* | 254 (24.8) | 372 (26.8) | 0.306 | 0.044 |
| Prev. Myocardial Inf. *N (%)* | 229 (16.5) | 161 (15.7) | 0.665 | 0.02 |
| Diabetes *N (%)* | 176 (17.2) | 245 (17.6) | 0.827 | 0.011 |
| Hypertension *N (%)* | 692 (67.6) | 995 (71.5) | 0.041 | 0.086 |
| Renal Failure *N (%)* | 213 (20.8) | 297 (21.4) | 0.784 | 0.013 |
| Sleep Apnea *N (%)* | 206 (14.8) | 154 (15.0) | 0.927 | 0.006 |
| Prev. PTCA *N (%)* | 335 (24.1) | 235 (22.9) | 0.548 | 0.027 |
| Prev. CABG *N (%)* | 132 ( 9.5) | 105 (10.3) | 0.579 | 0.026 |
| Previous Electroconversion *N (%)* | 355 (34.7) | 506 (36.4) | 0.410 | 0.036 |
| Prev. PVI *N (%)* | 279 (20.1) | 210 (20.5) | 0.825 | 0.011 |
| **Medication *N (%)*** | | | | |
| Antiplatelets | 53 ( 5.2) | 97 (7.0) | 0.085 | 0.075 |
| Aspirin | 171 (16.7) | 233 (16.8) | 1.000 | 0.001 |
| Statins | 529 (51.7) | 665 (47.8) | 0.067 | 0.077 |
| Diuretics | 459 (44.8) | 671 (48.2) | 0.105 | 0.068 |
| Betablockers | 726 (70.9) | 972 (69.9) | 0.619 | 0.022 |
| Digoxin | 39 ( 3.8) | 70 (5.0) | 0.183 | 0.060 |
| VKA | 414 (40.4) | 537 (38.6) | 0.387 | 0.037 |
| DOAC | 517 (50.5) | 713 (51.3) | 0.739 | 0.015 |
| **Implanted Device *N (%)*** | | | 0.909 | 0.051 |
| No device | 831 (81.2) | 1103 (79.3) | | |
| Loop recorder | 10 ( 1.0) | 14 (1.0) | | |
| PM | 121 (11.8) | 187 (13.4) | | |
| CRT | 12 ( 1.2) | 17 (1.2) | | |
| ICD | 31 ( 3.0) | 44 (3.2) | | |
| CRT-ICD | 19 ( 1.9) | 26 (1.9) | | |
| **Socioeconomic** | | | | |
| Mother tongue *N (%)* | | | 0.075 | 0.094 |
| German | 755 (73.7) | 1 075 (77.7) | | |
| French | 141 (13.8) | 163 (11.8) | | |
| Italian | 128 (12.5) | 145 (10.5) | | |
| Education *N (%)* | | | 0.086 | 0.091 |
| Basic | 139 (13.6) | 149 (10.7) | | |
| Middle | 505 (49.4) | 692 (49.9) | | |
| Advanced | 379 (37.0) | 547 (39.4) | | |
| Smoking *N (%)* | | | 0.882 | 0.021 |

| | | | | |
|---|---|---|---|---|
| Never | 453 (44.2) | 606 (43.6) | | |
| In the past | 495 (48.3) | 686 (49.3) | | |
| Active | 76 ( 7.4) | 99 (7.1) | | |
| Alcohol *mean (SD)* | 1.12 (1.53) | 1.04 (1.48) | 0.15 | 0.059 |
| **Greater Region** | | | <0.001 | 0.293 |
| Zurich | 56 ( 5.5) | 103 (7.4) | | |
| Lake Geneva Region | 289 (28.2) | 275 (19.9) | | |
| Espace Mittelland | 310 (30.3) | 568 (41.1) | | |
| Northwestern Switzerland | 67 ( 6.5) | 85 (6.1) | | |
| Eastern Switzerland | 125 (12.2) | 141 (10.2) | | |
| Southern Switzerland | 125 (12.2) | 124 (9.0) | | |
| Central Switzerland | 52 ( 5.1) | 87 (6.3) | | |

*Notes:* Alcohol in drinks per day. Abbreviations: AF: atrial fibrillation, CABG: coronary artery bypass grafting, $CHA_2DS_2$-VASc: risk of stroke (for non-valvular atrial fibrillation), CRT: Cardiac resynchronization therapy, DOAC: direct-acting oral anticoagulant, Dx: diagnosis, ICD: Implantable cardioverter defibrillator, IQR: interquartile range, PCG: pharmaceutical cost groups, PM: Pacemaker, Prev.: previous history of, PTCA: Percutaneous transluminal coronary angioplasty, PVI: Pulmonary vein isolation, SD: standard deviation, SMD: standardized mean difference, TIA: transient ischemic attack, VKA: vitamin K antagonist.

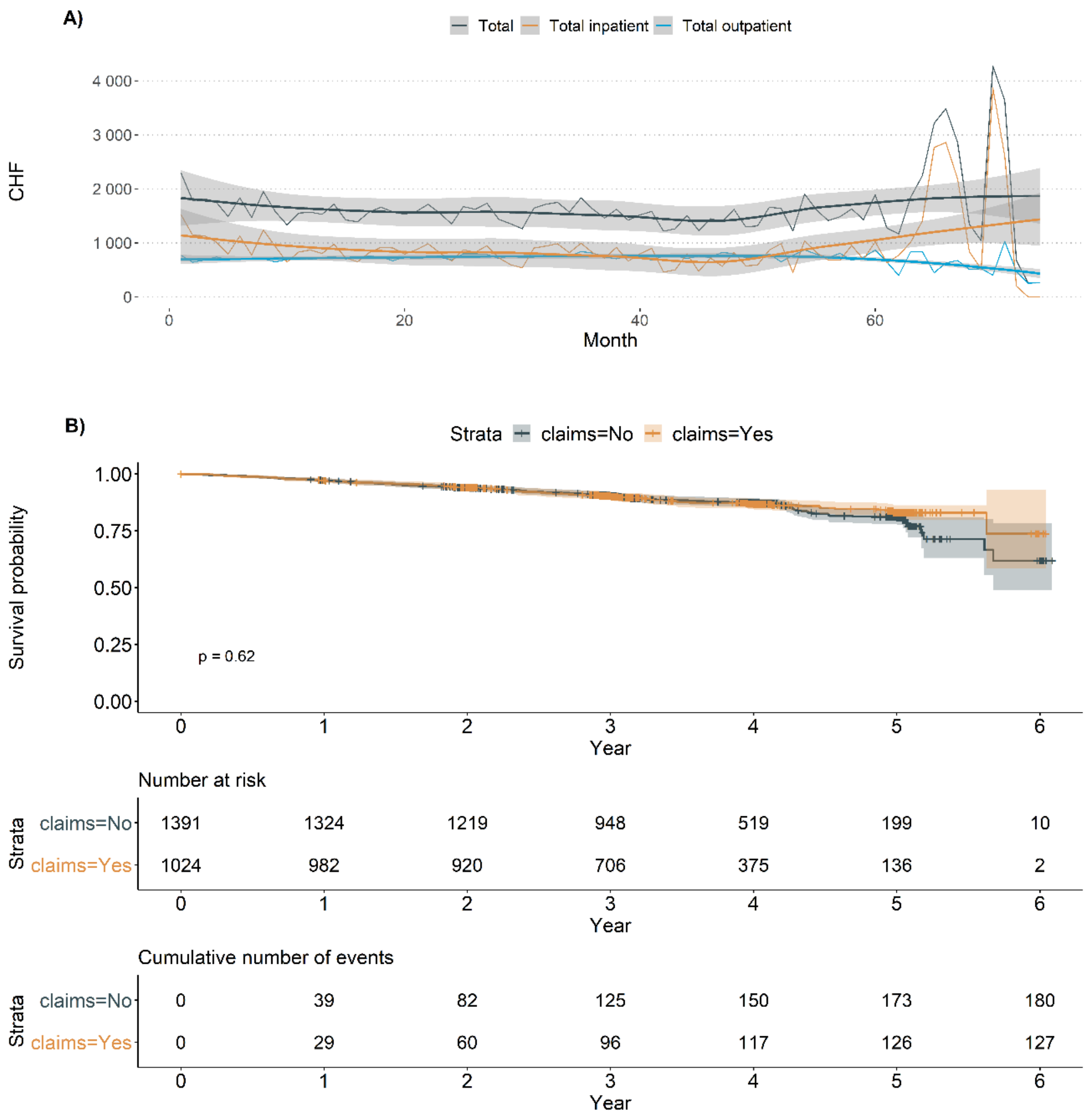

**Figure S2. Trajectory of monthly costs and Kaplan-Meier curve.** A) Temporal evolution of monthly costs for patients with claims data across all follow-ups. B) Overall survival and follow-up for all Swiss-AF patients: Kaplan-Meier curve, risk table, and cumulative number of events. *Notes:* median follow-up: 3.24 years; total patient-years of follow-up: 8 343.10.

**Table S2. Annual costs in Swiss Francs (CHF) by cost component.**

| Cost component | *Median [IQR]* | *Mean (SD)* |
|---|---|---|
| Total | 4 518 [825, 11 771] | 19 037 (59 998) |
| Total inpatient | 0 [0, 0] | 10 235 (56 327) |
| Total outpatient drugs | 508 [0, 2 956] | 2 495 (7 382) |
| Total outpatient without drugs | 2 282 [59, 7 225] | 6 307 (13 154) |
| Total AF-adj. | 400 [0, 3 213] | 5 679 (36 135) |
| Total AF-adj. inpatient | 0 [0, 0] | 3 458 (35 188) |
| Total AF-adj. outpatient drugs | 0 [0, 250] | 591 (1 392) |
| Total AF-adj. outpatient without drugs | 0 [0, 1 251] | 1 630 (6 899) |
| Total AF treatment | 226 [0, 2 773] | 4 078 (28 640) |
| Total stroke or TIA | 0 [0, 0] | 174 (9 124) |
| Total bleeding | 0 [0, 0] | 696 (17 462) |
| Total fall | 0 [0, 0] | 237 (4 434) |
| Total heart failure | 0 [0, 0] | 494 (8 469) |

*Notes:* Heart failure costs include inpatient services only. Abbreviations: adj.: adjudicated, AF: atrial fibrillation, IQR: interquartile range, SD: standard deviation, TIA: transient ischemic attack.

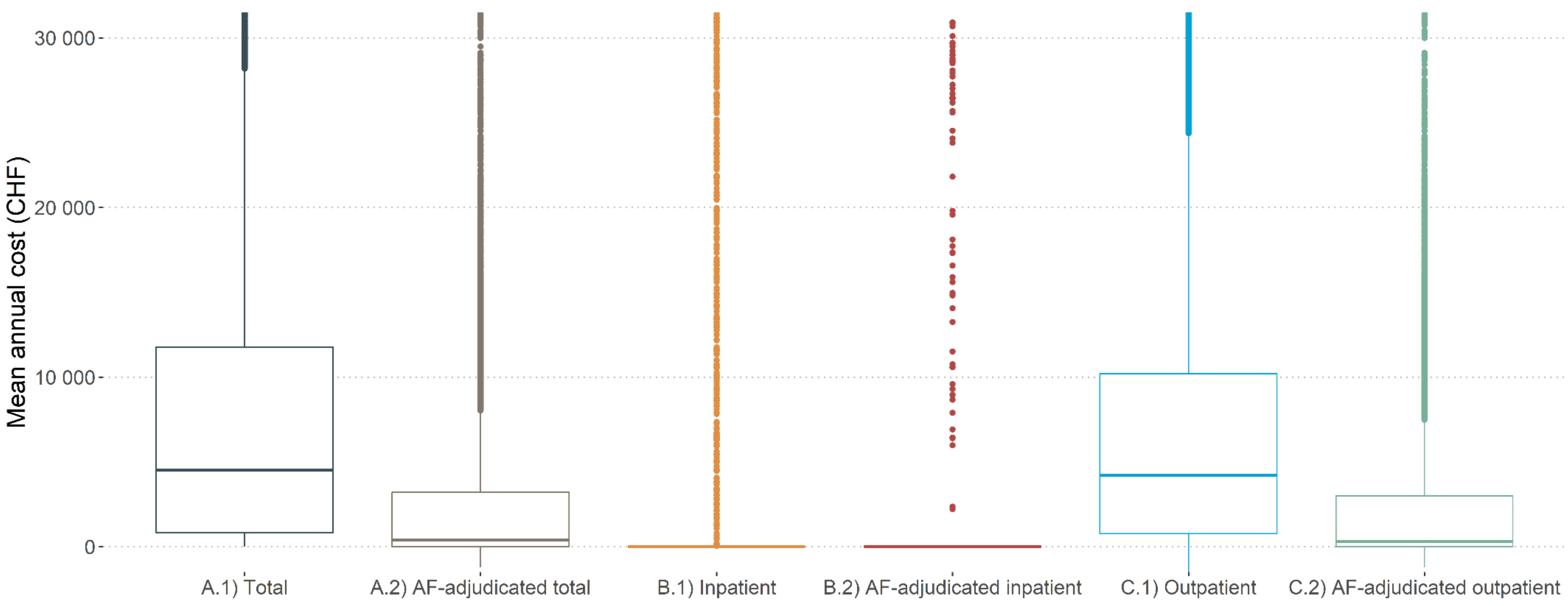

**Figure S3. Boxplot distribution of mean annual costs by cost outcome.**

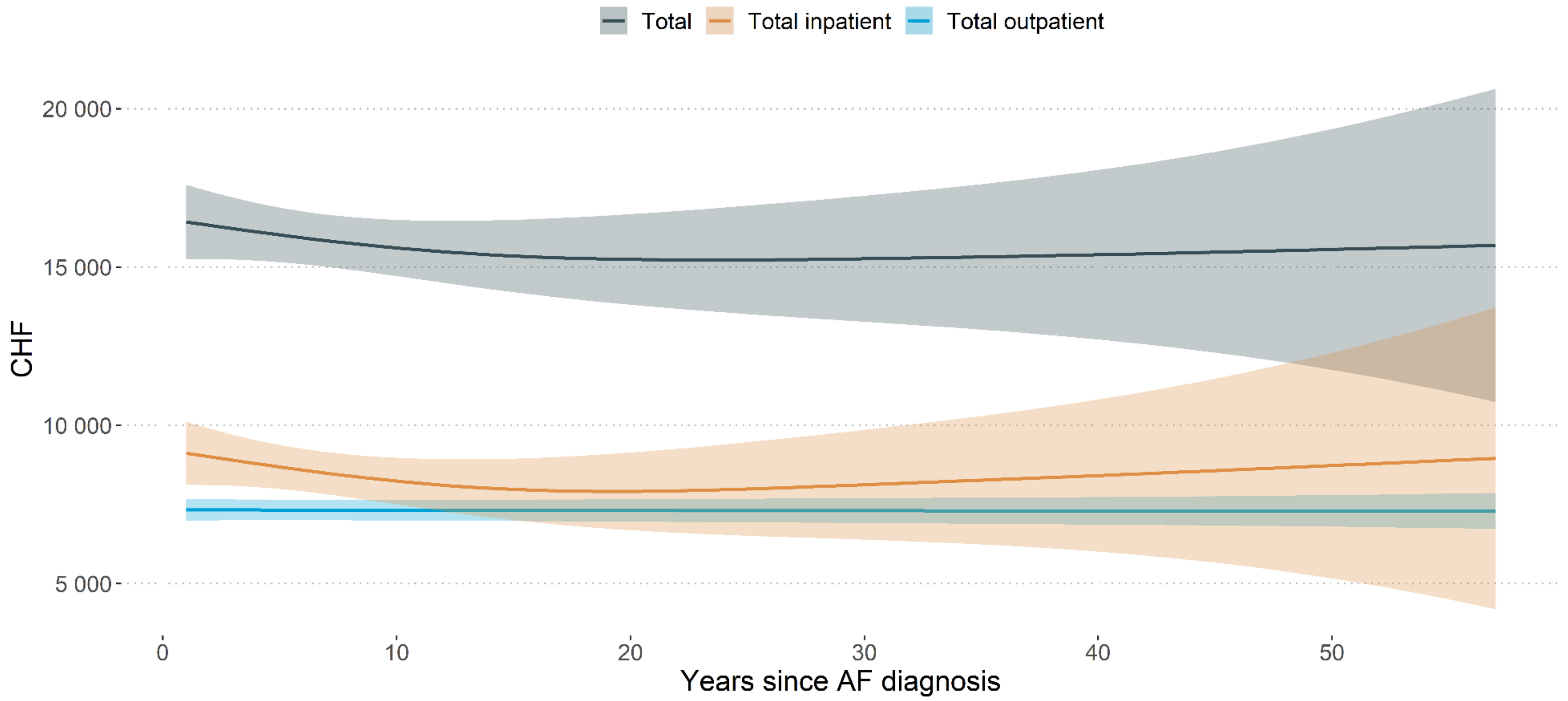

**Figure S4. Trajectory of mean annual costs since AF diagnosis.**

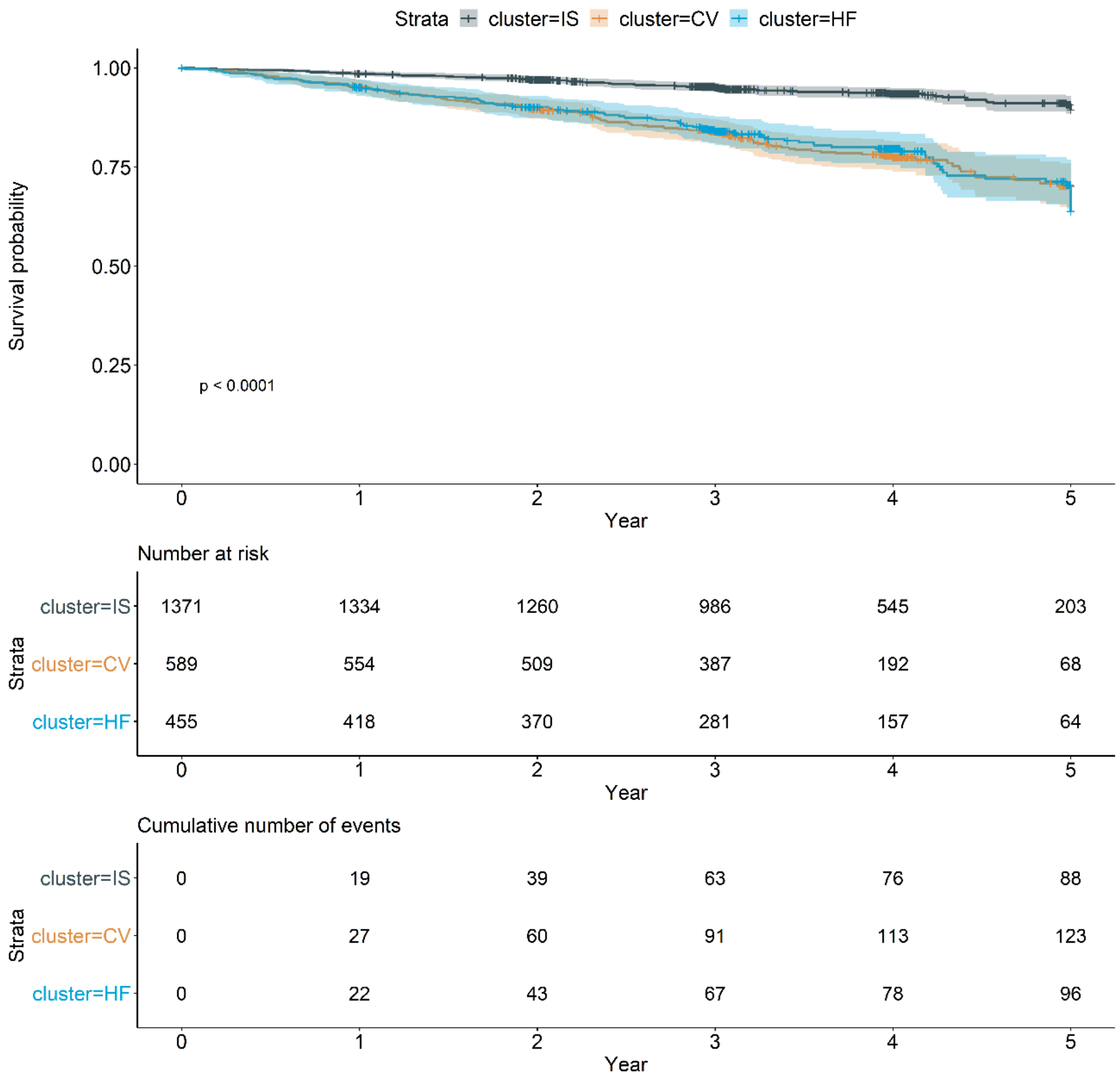

**Figure S4. Kaplan-Meier curve, risk table and cumulative number of events by patient cluster.** Cluster: CV = Cardiovascular Dominated, HF = Heart Failure Dominated, IS = Isolated Symptomatic AF.

**Table S3. Hazard Ratio of the Cox Regression.** Reference cluster = Isolated Symptomatic AF.

| | *HR* |
|---|---|
| Age | 1.09 [1.06, 1.11] |
| Time since Diagnosis | 1.01 [0.99, 1.03] |
| Cluster: Cardiovascular Dominated | 2.75 [1.79, 4.23] |
| Cluster: Heart Failure Dominated | 2.10 [1.30, 3.39] |
| Observations | 1 016 |
| $R^2$ | 0.081 |

**Table S4. Annual costs (CHF) of AF-adjudicated cost groups by patient cluster.**

| | **Cardiovascular Dominated** | | **Isolated Symptomatic AF** | | **Heart Failure Dominated** | |
|---|---|---|---|---|---|---|
| **Cost group** | *Median [IQR]* | *Mean (SD)* | *Median [IQR]* | *Mean (SD)* | *Median [IQR]* | *Mean (SD)* |
| Total AF treatment | 702 [0, 3 218] | 5 146 (36 440) | 0 [0, 2 423] | 3 637 (25 289) | 791 [0, 3 078] | 4 153 (27 545) |
| Total stroke / TIA | 0 [0, 0] | 170 (5 379) | 0 [0, 0] | 94 (7 010) | 0 [0, 0] | 447 (16 378) |
| Total bleeding | 0 [0, 0] | 1 512 (28 820) | 0 [0, 0] | 371 (10 593) | 0 [0, 0] | 713 (16 142) |
| Total fall | 0 [0, 0] | 252 (3 699) | 0 [0, 0] | 229 (4 911) | 0 [0, 0] | 247 (3 547) |
| Total heart failure | 0 [0, 0] | 1 104 (12 308) | 0 [0, 0] | 102 (3 513) | 0 [0, 0] | 1 005 (12 834) |

*Notes:* Heart failure costs include inpatient services only. Abbreviations: AF: atrial fibrillation, IQR: interquartile range, SD: standard deviation, TIA: transient ischemic attack.

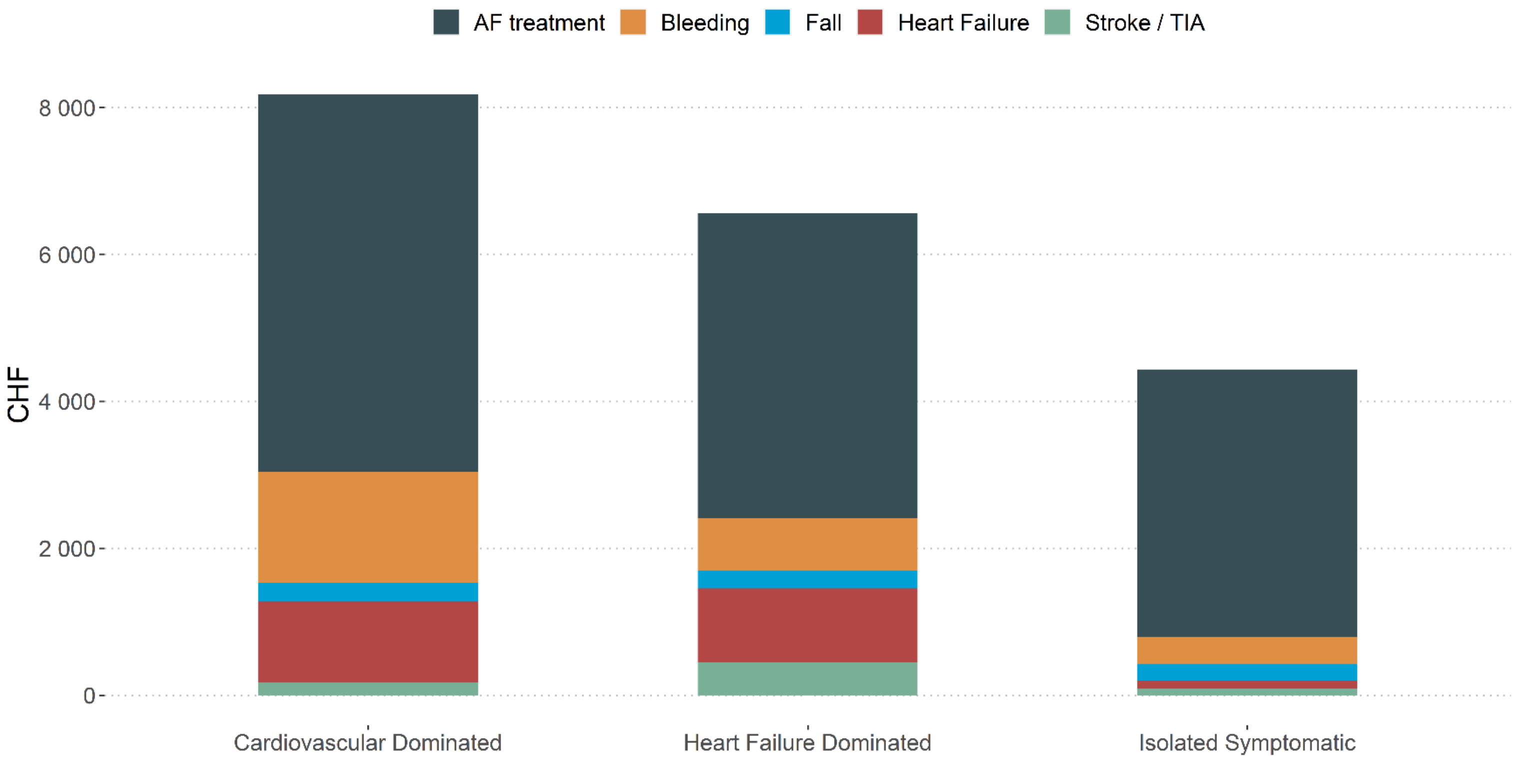

**Figure S5. Mean annual costs of adjudicated cost group by patient cluster.** Heart failure costs include inpatient services only. *Notes:* Abbreviations: AF: atrial fibrillation, TIA: transient ischemic attack.

**Table S5: Covariates associated with AF costs.**

*Introduction*

Complementary to the analysis in the main manuscript we aimed to evaluate the covariates associated with costs. For this, a multivariable regression-based analysis was run.

*Methods*

The covariates associated with costs were assessed using multivariable generalised linear models with quasi-poisson link distribution. Our model choice was motivated by the expected skewness of the outcome distributions[1,2] and by the straightforward interpretation of the exponentiated coefficients as rate ratios (RR), i.e. relative increases in cost.[3–6] Additional to the covariates listed in the manuscript we also considered further relevant comorbidities not collected by the Swiss-AF study. These were derived from the drug utilisation present in the claims data, using the pharmaceutical cost groups (PCGs) method[7]: acid related disorders, bone diseases, cancer, dementia, epilepsy, respiratory illness, rheumatic conditions, glaucoma, gout, iron deficiency, chronic pain, psychiatric diseases, use of antipsychotic drugs, thyroid disease, other rare diseases.

Several sensitivity analyses were carried out to assess the robustness of the results. As they are direct correlates of costs, covariates representing anticoagulation, other medication, treatment and implanted devices were left out to assess the isolated impact of medical history. In another sensitivity analysis, all but medical history predictors were included to assess treatment factors. Moreover, a gamma distribution was assumed instead of the quasi-poisson link to confirm appropriateness of the model choice. And last, the influence of the study centres as well as insurance characteristics on regression results were checked.

*Results*

Multivariable regression-based associations between covariates and AF-adjudicated costs are shown in **Figure S7**, while all numerical estimates are depicted in **Table S6**. The covariate patterns for AF-adjudicated total and AF-adjudicated inpatient costs were similar; however, associations with AF-adjudicated outpatient costs differed at some points. The significant effects for all three AF-adjudicated cost types include active smoking (RR 1.67; RR 1.77; RR 1.3), and AF symptoms (RR 1.43; RR 1.66; RR 1.08). An increase in AF-adjudicated total and AF-adjudicated inpatient costs arose also from permanent AF (RR 1.36; RR 1.61), previous myocardial infarction (RR 1.37; RR 1.66), and previous heart failure (RR 1.2; RR 1.43). In contrast, AF-adjudicated outpatient costs were significantly driven by the presence of any cardiac device (PM RR 1.22; ICD RR 1.76; CRT/CRT-ICD RR 1.62), and anticoagulation (VKA RR 1.45; DOAC RR 1.36). Two covariates were significantly associated with lower AF-adjudicated outpatient costs: history of stroke or TIA (RR 0.67), and being female (RR 0.77). The results of the sensitivity analyses can be found in **Figure S8**, and **Table S6-S8**. All results remained stable when insurance characteristics were additionally included.

*Discussion*

In our patient population, the factors most strongly associated with higher AF-adjudicated costs were active smoking and AF symptoms. Further associations were found to differ between AF-adjudicated inpatient versus outpatient costs. Estimates in the literature so far focussed on drivers of hospitalisation and total costs only.[8–11] Our estimates of a positive RR for permanent AF, BMI, and myocardial infarction are in line with the literature for AF-adjudicated inpatient and AF-adjudicated total costs. Unsurprisingly, medication like VKA or DOAC increased AF-adjudicated outpatient costs but had a small effect on AF-adjudicated inpatient costs. The association between presence of cardiac devices (pacemaker, ICD, and CRT) and AF-adjudicated outpatient costs was presumably due to routine controls and maintenance. Our estimates of sex-specific AF costs were inconclusive, in line with previous studies.[8,12]

Limitations for this analysis include the unfortunate case that we could not acquire insurance characteristics from one insurer and decided to only consider these in a sensitivity analysis. However, there was no indication of a distortion of our results. Another limitation lies therein, that some covariates were correlated and not all conceivable covariates were measured. This limited our ability to identify isolated effects. Even though different sensitivity analyses were run to check the robustness of the results, the observed associations should be interpreted cautiously.

*References*

1. Venables, W. N. & Ripley, B. D. *Modern Applied Statistics with S* . (Springer, 2002). doi:https://doi.org/10.1007/978-0-387-21706-2.
2. Miquel, L. *et al.* Alcohol, tobacco and health care costs: a population-wide cohort study (n = 606 947 patients) of current drinkers based on medical and administrative health records from Catalonia. *European Journal of Public Health* **28**, 674–680 (2018).
3. Mihaylova, B., Briggs, A., O'Hagan, A. & Thompson, S. G. Review of Statistical Methods for Analysing Healthcare Resources and Costs. *Health Economics* **20**, 897 (2011).
4. Austin, P., Ghali, W. A. & Tu, J. v. A comparison of several regression models for analysing cost of CABG surgery. *Statistics in Medicine* **22**, 2799–2815 (2003).

5. Bennell, M. C. *et al.* Identifying predictors of cumulative healthcare costs in incident atrial fibrillation: A population-based study. *J Am Heart Assoc* **4**, (2015).

6. Blough, D. K. & Ramsey, S. D. Using generalized linear models to assess medical care costs. *Health Services and Outcomes Research Methodology* **1**, 185–202 (2000).

7. Huber, C. A., Szucs, T. D., Rapold, R. & Reich, O. Identifying patients with chronic conditions using pharmacy data in Switzerland: an updated mapping approach to the classification of medications. *BMC Public Health* **13**, 1–10 (2013).

8. Bhat, A. *et al.* Drivers of hospitalization in atrial fibrillation: A contemporary review. *Heart Rhythm* **17**, 1991–1999 (2020).

9. Steinberg, B. A. *et al.* Drivers of hospitalization for patients with atrial fibrillation: Results from the Outcomes Registry for Better Informed Treatment of Atrial Fibrillation (ORBIT-AF). *American Heart Journal* **167**, 735-742.e2 (2014).

10. DeVore, A. D. *et al.* Hospitalizations in patients with atrial fibrillation: an analysis from ROCKET AF. *Europace* **18**, 1135–1142 (2016).

11. le Heuzey, J.-Y. *et al.* Cost of care distribution in atrial fibrillation patients: the COCAF study. *American Heart journal* **147**, 121–126 (2004).

12. Schnabel, R. B. *et al.* Gender differences in clinical presentation and 1-year outcomes in atrial fibrillation. *Heart* **103**, 1024–1030 (2017).

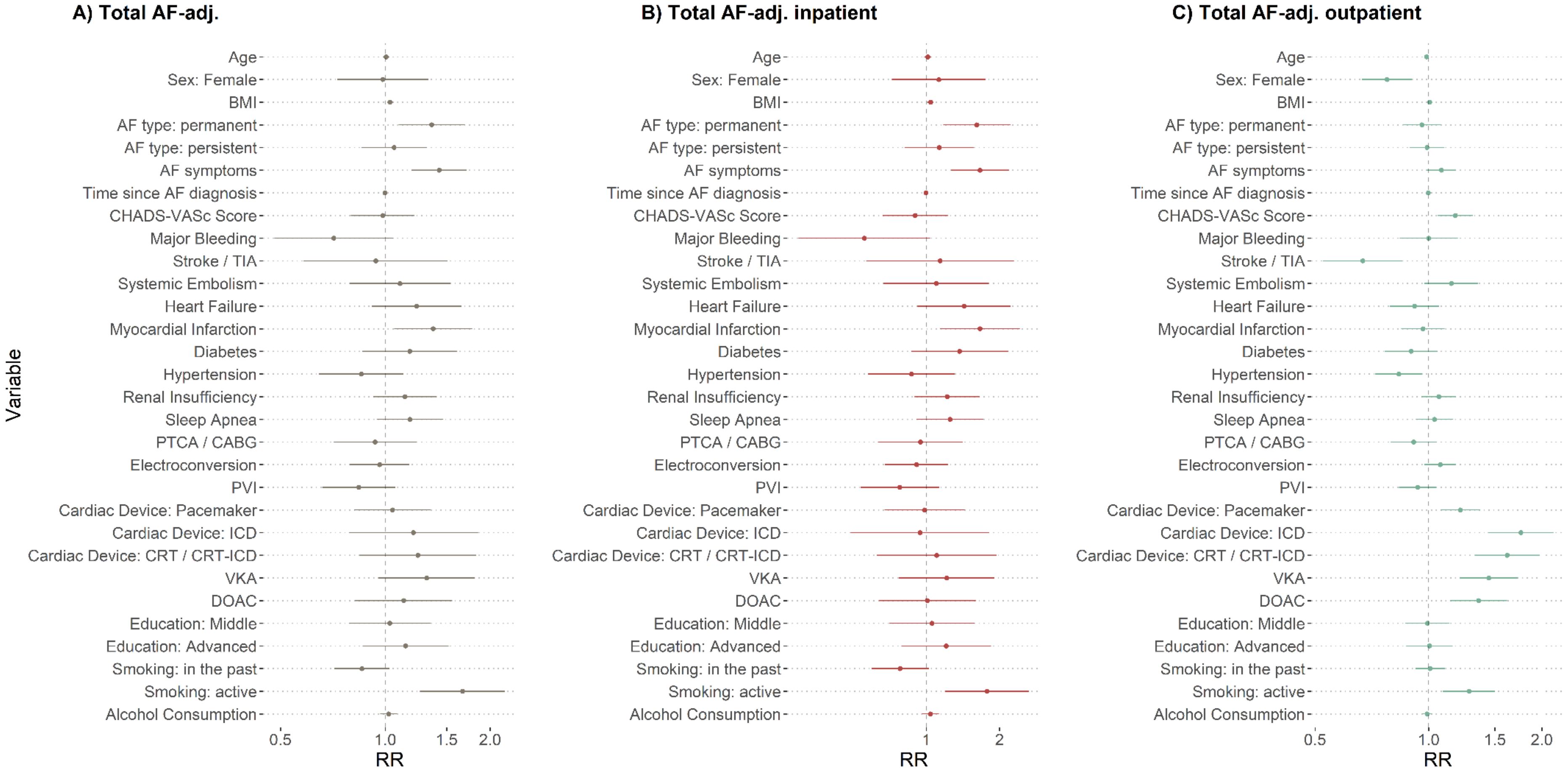

**Figure S7. Rate ratios (RR) and corresponding confidence intervals (CI) for the multivariable cost models.** A) RR and CI for factors associated with total AF-adjudicated costs. B) RR and CI for factors associated with total AF-adjudicated inpatient costs. C) RR and CI for factors associated with total AF-adjudicated outpatient costs.

*Notes:* Abbreviations: adj.: adjudicated, AF: atrial fibrillation, CABG: coronary artery bypass grafting, $CHA_2DS_2$-VASc: risk of stroke (for non-valvular atrial fibrillation), CRT: Cardiac resynchronization therapy, DOAC: direct-acting oral anticoagulant, ICD: Implantable cardioverter defibrillator, PCG: pharmaceutical cost groups, PTCA: Percutaneous transluminal coronary angioplasty, PVI: Pulmonary vein isolation, TIA: transient ischemic attack, VKA: vitamin K antagonist.

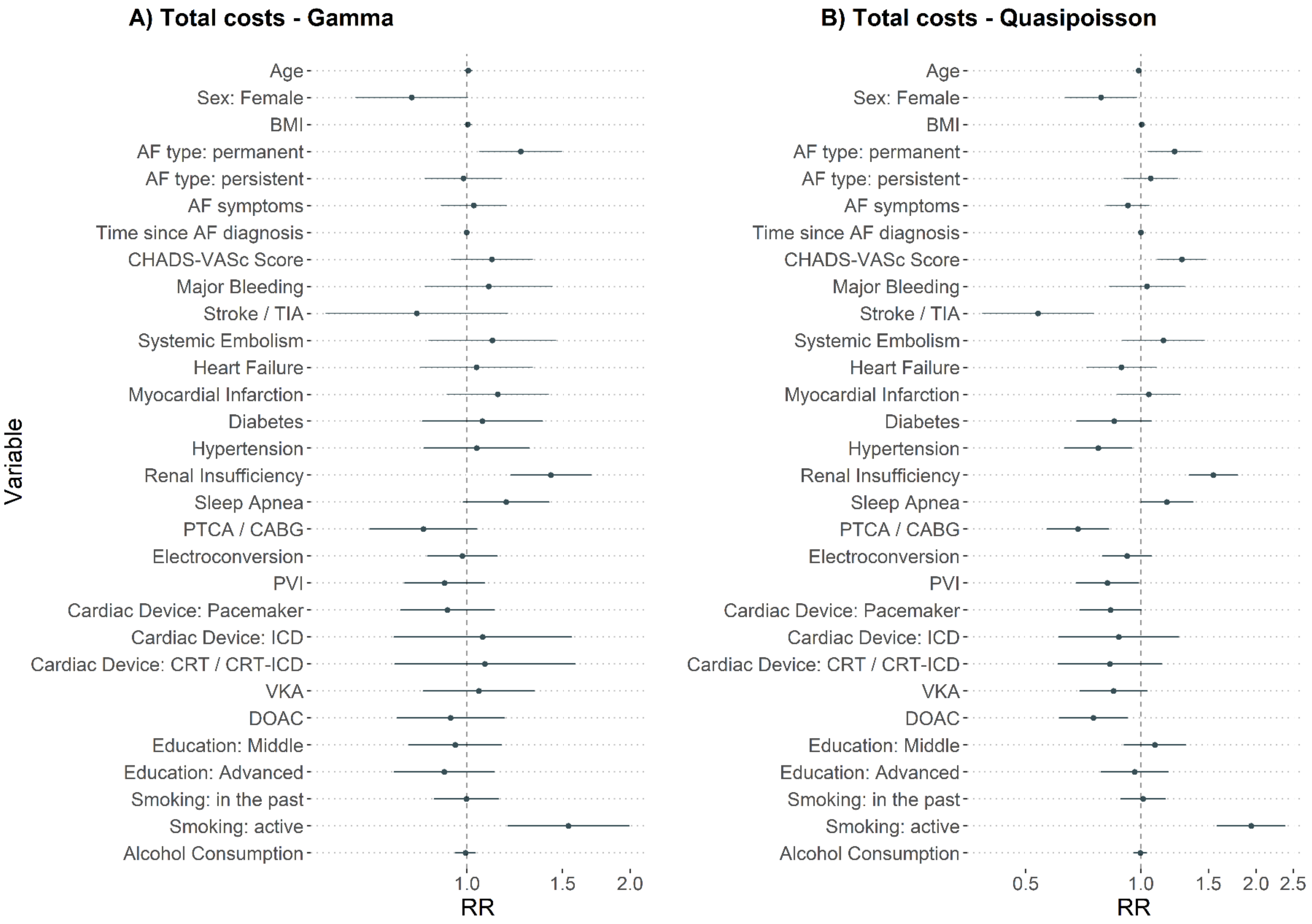

**Figure S8. Regression estimates for total costs using GLM Gamma (link = log) and Quasipoisson (link = log).**
*Notes:* Abbreviations: adj.: adjudicated, AF: atrial fibrillation, CABG: coronary artery bypass grafting, $CHA_2DS_2$-VASc: risk of stroke (for non-valvular atrial fibrillation), CRT: Cardiac resynchronization therapy, DOAC: direct-acting oral anticoagulant, ICD: Implantable cardioverter defibrillator, PCG: pharmaceutical cost groups, PTCA: Percutaneous transluminal coronary angioplasty, PVI: Pulmonary vein isolation, TIA: transient ischemic attack, VKA: vitamin K antagonist.

**Table S6. Estimates (RR) and confidence interval (95%) of multivariable cost models for different cost outcomes.**

| | *Dependent variable:* | | | | | |
|---|---|---|---|---|---|---|
| | **Total costs** | **Total AF-adj. costs** | **Total inpatient costs** | **Total AF-adj. inpatient costs** | **Total outpatient costs** | **Total AF-adj. outpatient costs** |
| Age | 0.99 [0.97, 1.00] | 1.01 [0.99, 1.03] | 0.99 [0.97, 1.01] | 1.01 [0.98, 1.04] | 0.99 [0.97, 1.00] | 0.99 [0.98, 1.00] |
| Sex: Female | 0.79 [0.64, 0.97] | 0.98 [0.73, 1.33] | 0.84 [0.63, 1.12] | 1.13 [0.72, 1.76] | 0.69 [0.57, 0.83] | 0.78 [0.67, 0.90] |
| BMI | 1.01 [0.99, 1.02] | [1.01, 1.05] | 1.00 [0.98, 1.02] | 1.04 [1.01, 1.07] | 1.01 [1.00, 1.02] | 1.01 [1.00, 1.02] |
| AF type: permanent | 1.22 [1.05, 1.43] | [1.09, 1.68] | 1.36 [1.10, 1.68] | 1.61 [1.18, 2.21] | 1.09 [0.95, 1.26] | 0.96 [0.86, 1.08] |
| Af type: persistent | 1.06 [0.90, 1.25] | 1.06 [0.85, 1.31] | 1.13 [0.90, 1.41] | 1.13 [0.82, 1.57] | 1.01 [0.88, 1.17] | 0.99 [0.89, 1.10] |
| AF symptoms | 0.92 [0.81, 1.05] | 1.43 [1.19, 1.71] | 0.93 [0.78, 1.11] | 1.66 [1.27, 2.18] | 0.99 [0.88, 1.11] | 1.08 [0.99, 1.18] |
| Years since AF diagnosis | 1.00 [0.99, 1.01] | 1.00 [0.99, 1.01] | 1.00 [0.99, 1.01] | 1.00 [0.98, 1.01] | 1.00 [0.99, 1.01] | 1.00 [0.99, 1.00] |
| $CHA_2DS_2$-VASc | 1.28 [1.10, 1.48] | 0.98 [0.80, 1.21] | 1.26 [1.03, 1.53] | 0.90 [0.66, 1.23] | 1.26 [1.11, 1.44] | 1.18 [1.06, 1.31] |
| Major Bleeding | 1.04 [0.83, 1.30] | 0.71 [0.48, 1.05] | 0.98 [0.72, 1.32] | 0.56 [0.30, 1.04] | 1.16 [0.95, 1.43] | 1.00 [0.84, 1.19] |
| Stroke / TIA | 0.54 [0.38, 0.75] | 0.94 [0.58, 1.51] | 0.54 [0.35, 0.85] | 1.14 [0.57, 2.29] | 0.59 [0.44, 0.80] | 0.67 [0.52, 0.85] |
| Systemic Embolism | 1.15 [0.89, 1.47] | 1.10 [0.79, 1.54] | 1.22 [0.88, 1.69] | 1.10 [0.67, 1.81] | 1.09 [0.87, 1.36] | 1.15 [0.98, 1.36] |
| Heart Failure | 0.89 [0.72, 1.10] | 1.23 [0.92, 1.66] | 1.01 [0.76, 1.34] | 1.43 [0.92, 2.22] | 0.70 [0.58, 0.85] | 0.92 [0.79, 1.06] |
| Myocardial Infarction | 1.05 [0.87, 1.27] | 1.37 [1.06, 1.77] | 1.11 [0.86, 1.43] | 1.66 [1.14, 2.42] | 1.02 [0.87, 1.21] | 0.97 [0.85, 1.10] |

| | | | | | | |
|---|---|---|---|---|---|---|
| Diabetes | 0.85 [0.68, 1.07] | 1.18 [0.86, 1.61] | 0.86 [0.64, 1.16] | 1.37 [0.87, 2.16] | 0.92 [0.75, 1.13] | 0.90 [0.77, 1.06] |
| Hypertension | 0.77 [0.63, 0.95] | 0.85 [0.65, 1.13] | 0.79 [0.60, 1.03] | 0.87 [0.58, 1.31] | 0.80 [0.67, 0.95] | 0.83 [0.72, 0.96] |
| Renal Insufficiency | 1.55 [1.34, 1.79] | 1.14 [0.92, 1.40] | 1.66 [1.36, 2.02] | 1.22 [0.89, 1.66] | 1.35 [1.19, 1.54] | 1.07 [0.96, 1.19] |
| Sleep Apnea | 1.17 [1.00, 1.37] | 1.18 [0.95, 1.46] | 1.24 [1.01, 1.53] | 1.25 [0.91, 1.72] | 1.12 [0.97, 1.29] | 1.04 [0.93, 1.16] |
| PTCA / CABG | 0.69 [0.57, 0.83] | 0.94 [0.71, 1.23] | 0.67 [0.52, 0.85] | 0.95 [0.64, 1.41] | 0.72 [0.60, 0.85] | 0.91 [0.79, 1.05] |
| Electroconversion | 0.92 [0.79, 1.07] | 0.96 [0.79, 1.17] | 0.93 [0.76, 1.14] | 0.91 [0.68, 1.23] | 0.92 [0.81, 1.05] | 1.07 [0.97, 1.19] |
| PVI | 0.82 [0.68, 0.99] | 0.84 [0.66, 1.07] | 0.71 [0.54, 0.93] | 0.78 [0.54, 1.13] | 0.98 [0.84, 1.14] | 0.94 [0.83, 1.05] |
| Cardiac Device: PM | 0.83 [0.69, 1.00] | 1.05 [0.82, 1.35] | 0.70 [0.54, 0.91] | 0.98 [0.67, 1.44] | 1.09 [0.93, 1.27] | 1.22 [1.07, 1.38] |
| Cardiac Device: ICD | 0.88 [0.61, 1.26] | 1.20 [0.79, 1.84] | 0.74 [0.45, 1.20] | 0.94 [0.49, 1.81] | 1.21 [0.88, 1.65] | 1.76 [1.44, 2.14] |
| Cardiac Device: CRT / CRT-ICD | 0.83 [0.61, 1.13] | 1.24 [0.84, 1.83] | 0.68 [0.44, 1.04] | 1.10 [0.63, 1.95] | 1.14 [0.87, 1.49] | 1.62 [1.33, 1.97] |
| VKA | 0.85 [0.69, 1.04] | 1.31 [0.95, 1.81] | 0.76 [0.58, 0.99] | 1.21 [0.77, 1.91] | 1.15 [0.94, 1.42] | 1.45 [1.21, 1.73] |
| DOAC | 0.75 [0.61, 0.92] | 1.13 [0.82, 1.56] | 0.64 [0.49, 0.84] | 1.01 [0.64, 1.60] | 1.08 [0.88, 1.33] | 1.36 [1.14, 1.62] |
| PCG acid related disorders | 1.15 [1.00, 1.33] | 1.24 [1.02, 1.50] | 1.11 [0.91, 1.35] | 1.20 [0.90, 1.61] | 1.23 [1.09, 1.40] | 1.32 [1.20, 1.45] |
| PCG bone diseases | 1.46 [1.16, 1.84] | 0.77 [0.50, 1.17] | 1.30 [0.93, 1.81] | 0.59 [0.30, 1.17] | 1.75 [1.45, 2.12] | 1.15 [0.96, 1.39] |

| | | | | | | |
|---|---|---|---|---|---|---|
| PCG cancer | 1.62 [1.26, 2.09] | 1.87 [1.30, 2.69] | 1.62 [1.14, 2.30] | 2.16 [1.28, 3.66] | 1.80 [1.46, 2.22] | 1.51 [1.25, 1.83] |
| PCG dementia | 0.99 [0.68, 1.42] | 1.14 [0.69, 1.88] | 0.86 [0.50, 1.47] | 0.99 [0.45, 2.20] | 1.19 [0.89, 1.58] | 1.32 [1.05, 1.65] |
| PCG epilepsy | 1.64 [1.35, 2.00] | 1.88 [1.44, 2.47] | 1.83 [1.41, 2.38] | 2.16 [1.47, 3.16] | 1.35 [1.13, 1.62] | 1.18 [1.01, 1.38] |
| PCG glaucoma | 0.99 [0.82, 1.21] | 0.88 [0.66, 1.16] | 0.86 [0.65, 1.14] | 0.79 [0.52, 1.22] | 1.19 [1.01, 1.39] | 1.01 [0.88, 1.16] |
| PCG gout | 0.92 [0.76, 1.11] | 0.89 [0.67, 1.17] | 0.89 [0.69, 1.15] | 0.85 [0.57, 1.28] | 1.01 [0.85, 1.19] | 0.97 [0.84, 1.11] |
| PCG iron deficiency | 1.39 [1.14, 1.70] | 1.08 [0.80, 1.46] | 1.32 [1.00, 1.74] | 1.07 [0.69, 1.67] | 1.56 [1.32, 1.84] | 1.15 [0.99, 1.33] |
| PCG pain | 1.04 [0.90, 1.20] | 1.01 [0.83, 1.23] | 0.97 [0.80, 1.18] | 0.97 [0.73, 1.30] | 1.17 [1.03, 1.32] | 1.07 [0.97, 1.18] |
| PCG psychiatric | 1.11 [0.96, 1.27] | 1.12 [0.92, 1.36] | 1.05 [0.86, 1.28] | 1.13 [0.84, 1.50] | 1.16 [1.03, 1.30] | 1.12 [1.02, 1.23] |
| PCG antipsychotic | 0.81 [0.52, 1.26] | 0.65 [0.34, 1.26] | 0.90 [0.50, 1.63] | 0.54 [0.20, 1.49] | 0.76 [0.52, 1.11] | 0.85 [0.62, 1.18] |
| PCG respiratory | 1.00 [0.85, 1.18] | 0.88 [0.70, 1.12] | 0.91 [0.73, 1.14] | 0.85 [0.60, 1.21] | 1.19 [1.04, 1.36] | 0.99 [0.88, 1.11] |
| PCG rheumatic conditions | 1.05 [0.91, 1.21] | 0.94 [0.77, 1.14] | 1.07 [0.88, 1.30] | 0.89 [0.67, 1.19] | 1.04 [0.92, 1.17] | 1.02 [0.93, 1.13] |
| PCG thyroid disorders | 1.22 [1.00, 1.50] | 1.41 [1.08, 1.84] | 1.27 [0.96, 1.68] | 1.64 [1.11, 2.40] | 1.21 [1.01, 1.44] | 1.12 [0.97, 1.28] |
| PCG sparse | 1.08 [0.80, 1.47] | 0.56 [0.32, 0.96] | 0.89 [0.56, 1.40] | 0.49 [0.21, 1.15] | 1.29 [1.01, 1.65] | 0.75 [0.58, 0.97] |

| | | | | | | |
|---|---|---|---|---|---|---|
| Education: Middle | 1.09 [0.90, 1.31] | 1.03 [0.79, 1.34] | 1.09 [0.85, 1.40] | 1.06 [0.71, 1.57] | 1.07 [0.91, 1.27] | 0.99 [0.87, 1.13] |
| Education: Advanced | 0.96 [0.79, 1.18] | 1.14 [0.86, 1.52] | 0.88 [0.67, 1.16] | 1.21 [0.79, 1.84] | 1.08 [0.90, 1.29] | 1.01 [0.87, 1.16] |
| Smoking: In the past | 1.01 [0.88, 1.16] | 0.86 [0.71, 1.03] | 1.06 [0.88, 1.27] | 0.78 [0.59, 1.03] | 0.92 [0.82, 1.04] | 1.01 [0.92, 1.11] |
| Smoking: Active | 1.94 [1.58, 2.39] | 1.67 [1.26, 2.20] | 2.44 [1.87, 3.18] | 1.78 [1.19, 2.64] | 1.15 [0.94, 1.42] | 1.28 [1.09, 1.50] |
| Alcohol | 1.00 [0.96, 1.04] | 1.02 [0.97, 1.08] | 1.00 [0.95, 1.06] | 1.04 [0.96, 1.12] | 1.00 [0.96, 1.03] | 0.99 [0.96, 1.02] |
| Greater Region: Lake Geneva | 1.17 [0.83, 1.64] | 0.85 [0.54, 1.32] | 1.14 [0.70, 1.88] | 0.82 [0.41, 1.65] | 1.16 [0.88, 1.53] | 0.98 [0.80, 1.19] |
| Greater Region: Espace Mittelland | 1.32 [1.05, 1.65] | 1.16 [0.88, 1.54] | 1.50 [1.09, 2.07] | 1.35 [0.88, 2.08] | 1.09 [0.91, 1.31] | 0.92 [0.80, 1.05] |
| Greater Region: Northwestern Switzerland | 1.67 [1.34, 2.07] | 1.14 [0.86, 1.51] | 2.13 [1.56, 2.91] | 1.40 [0.91, 2.15] | 1.13 [0.95, 1.35] | 0.86 [0.75, 0.98] |
| Greater Region: Eastern Switzerland | 1.12 [0.80, 1.57] | 0.84 [0.54, 1.31] | 1.27 [0.79, 2.03] | 0.90 [0.46, 1.78] | 0.98 [0.74, 1.29] | 0.80 [0.65, 0.98] |
| Greater Region: Southern Switzerland | 1.12 [0.86, 1.47] | 0.70 [0.49, 1.01] | 1.11 [0.75, 1.64] | 0.68 [0.39, 1.20] | 1.15 [0.93, 1.44] | 0.78 [0.66, 0.93] |
| Greater Region: Central Switzerland | 0.98 [0.65, 1.48] | 0.97 [0.60, 1.56] | 1.26 [0.71, 2.23] | 1.11 [0.53, 2.31] | 0.73 [0.51, 1.02] | 0.79 [0.63, 0.99] |
| Observations | 1013 | 1013 | 1013 | 1013 | 1013 | 1013 |

*Notes:* Alcohol in drinks per day. Abbreviations: AF: atrial fibrillation, CABG: coronary artery bypass grafting, $CHA_2DS_2$-VASc: risk of stroke (for non-vavluar atrial fibrillation), CRT: Cardiac resynchronization therapy, DOAC: direct-acting oral anticoagulant, Dx: diagnosis, ICD: Implantable cardioverter defibrillator, PCG: pharmaceutical cost groups, PM: Pacemaker, PTCA: Percutaneous transluminal coronary angioplasty, PVI: Pulmonary vein isolation, TIA: transient ischemic attack, VKA: vitamin K antagonist.

**Table S7. Regression estimates for total costs using GLM Gamma (link = log) and Quasipoisson (link = log).**

| | Total costs | |
|---|---|---|
| | *Gamma* | *Quasipoisson* |
| Age | 1.01 [0.99, 1.02] | 0.99 [0.97, 1.00] |
| Sex: Female | 0.79 [0.62, 1.00] | 0.79 [0.64, 0.97] |
| BMI | 1.00 [0.99, 1.02] | 1.01 [0.99, 1.02] |
| AF type: permanent | 1.26 [1.06, 1.50] | 1.22 [1.05, 1.43] |
| AF type: persistent | 0.99 [0.84, 1.16] | 1.06 [0.90, 1.25] |
| AF symptoms | 1.03 [0.90, 1.18] | 0.92 [0.81, 1.05] |
| Years since AF diagnosis | 1.00 [0.99, 1.01] | 1.00 [0.99, 1.01] |
| $CHA_2DS_2$-VASc | 1.11 [0.94, 1.32] | 1.28 [1.10, 1.48] |
| Major Bleeding | 1.10 [0.84, 1.43] | 1.04 [0.83, 1.30] |
| Stroke / TIA | 0.81 [0.55, 1.19] | 0.54 [0.38, 0.75] |
| Systemic Embolism | 1.11 [0.85, 1.46] | 1.15 [0.89, 1.47] |
| Heart Failure | 1.04 [0.82, 1.32] | 0.89 [0.72, 1.10] |
| Myocardial Infarction | 1.14 [0.92, 1.41] | 1.05 [0.87, 1.27] |
| Diabetes | 1.07 [0.83, 1.38] | 0.85 [0.68, 1.07] |
| Hypertension | 1.04 [0.83, 1.31] | 0.77 [0.63, 0.95] |
| Renal Insufficiency | 1.43 [1.20, 1.69] | 1.55 [1.34, 1.79] |
| Sleep Apnea | 1.18 [0.98, 1.42] | 1.17 [1.00, 1.37] |
| PTCA / CABG | 0.83 [0.66, 1.04] | 0.69 [0.57, 0.83] |
| Electroconversion | 0.98 [0.84, 1.14] | 0.92 [0.79, 1.07] |
| PVI | 0.91 [0.77, 1.08] | 0.82 [0.68, 0.99] |
| Cardiac Device: PM | 0.92 [0.75, 1.12] | 0.83 [0.69, 1.00] |
| Cardiac Device: ICD | 1.07 [0.73, 1.56] | 0.88 [0.61, 1.26] |
| Cardiac Device: CRT / CRT-ICD | 1.08 [0.73, 1.58] | 0.83 [0.61, 1.13] |
| VKA | 1.05 [0.83, 1.33] | 0.85 [0.69, 1.04] |
| DOAC | 0.93 [0.74, 1.17] | 0.75 [0.61, 0.92] |
| Education: Middle | 0.95 [0.78, 1.16] | 1.09 [0.90, 1.31] |
| Education: Advanced | 0.91 [0.73, 1.12] | 0.96 [0.79, 1.18] |
| Smoking: In the past | 1.00 [0.87, 1.14] | 1.01 [0.88, 1.16] |
| Smoking: Active | 1.54 [1.19, 1.99] | 1.94 [1.58, 2.39] |
| Alcohol Consumption | 0.99 [0.95, 1.04] | 1.00 [0.96, 1.04] |

*Notes:* Alcohol in drinks per day. Abbreviations: AF: atrial fibrillation, CABG: coronary artery bypass grafting, $CHA_2DS_2$-VASc: risk of stroke (for non-valvular atrial fibrillation), CRT: Cardiac resynchronization therapy, Dx: diagnosis, ICD: Implantable cardioverter defibrillator, DOAC: direct-acting oral anticoagulant, PCG: pharmaceutical cost groups, PM: Pacemaker, PTCA: Percutaneous transluminal coronary angioplasty, PVI: Pulmonary vein isolation, TIA: transient ischemic attack, VKA: vitamin K antagonist.

**Table S8. Comparison of three multivariable cost models for total AF-adjudicated costs.**

| | Total AF-adjudicated costs | | |
|---|---|---|---|
| | *Main* | *Medical History* | *Treatment* |
| Observation time | 0.98 [0.98, 0.99] | 0.98 [0.98, 0.99] | 0.98 [0.98, 0.99] |
| Age | 1.01 [0.99, 1.03] | 1.01 [0.99, 1.03] | 1.00 [0.99, 1.01] |
| Sex: Female | 0.98 [0.73, 1.33] | 1.00 [0.76, 1.30] | 0.90 [0.72, 1.12] |
| BMI | 1.03 [1.01, 1.05] | 1.03 [1.01, 1.05] | 1.03 [1.01, 1.04] |
| AF type: permanent | 1.36 [1.09, 1.68] | 1.45 [1.18, 1.79] | 1.29 [1.04, 1.61] |
| Af type: persistent | 1.06 [0.85, 1.31] | 1.07 [0.88, 1.30] | 1.06 [0.85, 1.31] |
| AF symptoms | 1.43 [1.19, 1.71] | 1.42 [1.18, 1.70] | 1.52 [1.27, 1.82] |
| Years since AF diagnosis | 1.00 [0.99, 1.01] | 1.00 [0.99, 1.01] | 1.00 [0.99, 1.01] |
| $CHA_2DS_2$-VASc | 0.98 [0.80, 1.21] | 0.96 [0.81, 1.14] | - |
| Major Bleeding | 0.71 [0.48, 1.05] | 0.70 [0.47, 1.03] | - |
| Stroke / TIA | 0.94 [0.58, 1.51] | 0.97 [0.65, 1.45] | - |
| Systemic Embolism | 1.10 [0.79, 1.54] | 1.11 [0.80, 1.55] | - |
| Heart Failure | 1.23 [0.92, 1.66] | 1.32 [1.02, 1.72] | - |
| Myocardial Infarction | 1.37 [1.06, 1.77] | 1.40 [1.10, 1.79] | - |
| Diabetes | 1.18 [0.86, 1.61] | 1.23 [0.93, 1.63] | - |
| Hypertension | 0.85 [0.65, 1.13] | 0.85 [0.66, 1.10] | - |
| Renal Insufficiency | 1.14 [0.92, 1.40] | 1.17 [0.95, 1.44] | - |
| Sleep Apnea | 1.18 [0.95, 1.46] | 1.17 [0.94, 1.45] | - |
| PTCA / CABG | 0.94 [0.71, 1.23] | - | 0.99 [0.80, 1.23] |
| Electroconversion | 0.96 [0.79, 1.17] | - | 0.98 [0.80, 1.19] |
| PVI | 0.84 [0.66, 1.07] | - | 0.85 [0.67, 1.08] |
| Cardiac Device: PM | 1.05 [0.82, 1.35] | - | 1.03 [0.80, 1.33] |
| Cardiac Device: ICD | 1.20 [0.79, 1.84] | - | 1.28 [0.84, 1.95] |
| Cardiac Device: CRT / CRT-ICD | 1.24 [0.84, 1.83] | - | 1.46 [1.00, 2.11] |
| Antiplatelet | - | - | 1.54 [1.08, 2.19] |
| Aspirin | - | - | 0.96 [0.74, 1.24] |
| Statin | - | - | 1.11 [0.92, 1.33] |
| Diuretics | - | - | 1.52 [1.27, 1.83] |
| Betablocker | - | - | 0.96 [0.79, 1.16] |
| Digoxin | - | - | 0.69 [0.43, 1.09] |
| VKA | 1.31 [0.95, 1.81] | - | 1.30 [0.91, 1.84] |
| DOAC | 1.13 [0.82, 1.56] | - | 1.12 [0.79, 1.59] |
| PCG acid related disorders | 1.24 [1.02, 1.50] | 1.23 [1.01, 1.49] | 1.22 [1.00, 1.49] |
| PCG bone diseases | 0.77 [0.50, 1.17] | 0.79 [0.52, 1.20] | 0.71 [0.47, 1.09] |
| PCG cancer | 1.87 [1.30, 2.69] | 1.89 [1.32, 2.72] | 1.80 [1.25, 2.59] |
| PCG dementia | 1.14 [0.69, 1.88] | 1.14 [0.69, 1.87] | 1.06 [0.64, 1.75] |
| PCG epilepsy | 1.88 [1.44, 2.47] | 1.89 [1.44, 2.47] | 1.84 [1.41, 2.41] |
| PCG glaucoma | 0.88 [0.66, 1.16] | 0.89 [0.67, 1.18] | 0.89 [0.67, 1.17] |

| | | | |
|---|---|---|---|
| PCG gout | 0.89 [0.67, 1.17] | 0.90 [0.68, 1.17] | 0.90 [0.68, 1.17] |
| PCG iron deficiency | 1.08 [0.80, 1.46] | 1.12 [0.84, 1.51] | 1.10 [0.81, 1.48] |
| PCG pain | 1.01 [0.83, 1.23] | 1.00 [0.83, 1.22] | 1.07 [0.88, 1.29] |
| PCG psychiatric | 1.12 [0.92, 1.36] | 1.12 [0.92, 1.36] | 1.09 [0.90, 1.32] |
| PCG antipsychotic | 0.65 [0.34, 1.26] | 0.71 [0.37, 1.34] | 0.62 [0.32, 1.20] |
| PCG respiratory | 0.88 [0.70, 1.12] | 0.88 [0.70, 1.10] | 0.88 [0.70, 1.12] |
| PCG rheumatic conditions | 0.94 [0.77, 1.14] | 0.92 [0.76, 1.11] | 0.96 [0.79, 1.16] |
| PCG thyroid disorders | 1.41 [1.08, 1.84] | 1.43 [1.10, 1.87] | 1.39 [1.07, 1.81] |
| PCG sparse | 0.56 [0.32, 0.96] | 0.54 [0.31, 0.94] | 0.59 [0.34, 1.03] |
| Education: Middle | 1.03 [0.79, 1.34] | 1.02 [0.78, 1.32] | 1.04 [0.80, 1.36] |
| Education: Advanced | 1.14 [0.86, 1.52] | 1.09 [0.82, 1.44] | 1.18 [0.89, 1.57] |
| Smoking: In the past | 0.86 [0.71, 1.03] | 0.85 [0.71, 1.02] | 0.82 [0.69, 0.99] |
| Smoking: Active | 1.67 [1.26, 2.20] | 1.65 [1.25, 2.18] | 1.58 [1.19, 2.10] |
| Alcohol | 1.02 [0.97, 1.08] | 1.03 [0.97, 1.08] | 1.01 [0.96, 1.07] |
| Greater Region: Lake Geneva | 0.85 [0.54, 1.32] | 0.90 [0.58, 1.40] | 0.92 [0.59, 1.43] |
| Greater Region: Espace Mittelland | 1.16 [0.88, 1.54] | 1.22 [0.92, 1.61] | 1.19 [0.90, 1.58] |
| Greater Region: Northwestern Switzerland | 1.14 [0.86, 1.51] | 1.21 [0.92, 1.60] | 1.16 [0.88, 1.54] |
| Greater Region: Eastern Switzerland | 0.84 [0.54, 1.31] | 0.87 [0.56, 1.35] | 0.89 [0.57, 1.38] |
| Greater Region: Southern Switzerland | 0.70 [0.49, 1.01] | 0.73 [0.51, 1.06] | 0.68 [0.48, 0.98] |
| Greater Region: Central Switzerland | 0.97 [0.60, 1.56] | 0.96 [0.60, 1.55] | 0.86 [0.53, 1.39] |
| Observations | 1013 | 1013 | 1013 |

*Notes:* Alcohol in drinks per day. Abbreviations: AF: atrial fibrillation, CABG: coronary artery bypass grafting, $CHA_2DS_2$-VASc: risk of stroke (for non-valvular atrial fibrillation), CRT: Cardiac resynchronization therapy, DOAC: direct-acting oral anticoagulant, Dx: diagnosis, ICD: Implantable cardioverter defibrillator, PCG: pharmaceutical cost groups, PM: Pacemaker, PTCA: Percutaneous transluminal coronary angioplasty, PVI: Pulmonary vein isolation, TIA: transient ischemic attack, VKA: vitamin K antagonist.

**Swiss-AF investigators**

*University Hospital Basel and Basel University:* Stefanie Aeschbacher, Katalin Bhend, Steffen Blum, Leo Bonati, David Conen, Ceylan Eken, Urs Fischer, Corinne Girroy, Elisa Hennings, Elena Herber, Vasco Iten, Philipp Krisai, Michael Kühne, Maurin Lampart, Mirko Lischer, Nina Mäder, Christine Meyer-Zürn, Pascal Meyre, Andreas U. Monsch, Luke Mosher, Christian Müller, Stefan Osswald, Rebecca E. Paladini, Anne Springer, Christian Sticherling, Thomas Szucs, Gian Völlmin.
Principal Investigator: Stefan Osswald; Local Principal Investigator: Michael Kühne

*University Hospital Bern:* Faculty: Drahomir Aujesky, Juerg Fuhrer, Laurent Roten, Simon Jung, Heinrich Mattle; Research fellows: Seraina Netzer, Luise Adam, Carole Elodie Aubert, Martin Feller, Axel Loewe, Elisavet Moutzouri, Claudio Schneider;
Study nurses: Tanja Flückiger, Cindy Groen, Lukas Ehrsam, Sven Hellrigl, Alexandra Nuoffer, Damiana Rakovic, Nathalie Schwab, Rylana Wenger, Tu Hanh Zarrabi Saffari.
Local Principal Investigator: Nicolas Rodondi, Tobias Reichlin

*Stadtspital Triemli Zurich:* Christopher Beynon, Roger Dillier, Michèle Deubelbeiss, Franz Eberli, Christine Franzini, Isabel Juchli, Claudia Liedtke, Samira Murugiah, Jacqueline Nadler, Thayze Obst, Jasmin Roth, Fiona Schlomowitsch, Xiaoye Schneider, Katrin Studerus, Noreen Tynan, Dominik Weishaupt.
Local Principal Investigator: Andreas Müller

*Kantonspital Baden:* Simone Fontana, Corinne Friedli, Silke Kuest, Karin Scheuch, Denise Hischier, Nicole Bonetti, Alexandra Grau, Jonas Villinger, Eva Laube, Philipp Baumgartner, Mark Filipovic, Marcel Frick, Giulia Montrasio, Stefanie Leuenberger, Franziska Rutz.
Local Principal Investigator: Jürg-Hans Beer

*Cardiocentro Lugano:* Angelo Auricchio, Adriana Anesini, Cristina Camporini, Giulio Conte, Maria Luce Caputo, Francois Regoli.
Local Principal Investigator: Tiziano Moccetti

*Kantonsspital St. Gallen:* Roman Brenner, David Altmann, Michaela Gemperle.
Local Principal Investigator: Peter Ammann

*Hôpital Cantonal Fribourg:* Mathieu Firmann, Sandrine Foucras, Martine Rime.
Local Principal Investigator: Daniel Hayoz

*Luzerner Kantonsspital:* Benjamin Berte, Kathrin Bühler, Virgina Justi, Frauke Kellner-Weldon, Brigitta Mehmann, Sonja Meier, Myriam Roth, Andrea Ruckli-Kaeppeli, Ian Russi, Kai Schmidt, Mabelle Young, Melanie Zbinden.
Local Principal Investigator: Richard Kobza

*Ente Ospedaliero Cantonale Lugano:* Elia Rigamonti, Carlo Cereda, Alessandro Cianfoni, Maria Luisa De Perna, Jane Frangi-Kultalahti, Patrizia Assunta Mayer Melchiorre, Anica Pin,Tatiana Terrot, Luisa Vicari.
Local Principal Investigator: Giorgio Moschovitis.

*University Hospital Geneva:* Georg Ehret, Hervé Gallet, Elise Guillermet, Francois Lazeyras, Karl-Olof Lovblad, Patrick Perret, Philippe Tavel, Cheryl Teres.
Local Principal Investigator: Dipen Shah

*University Hospital Lausanne:* Nathalie Lauriers, Marie Méan, Sandrine Salzmann, Jürg Schläpfer.
Local Principal Investigator: Alessandra Pia Porretta

*Bürgerspital Solothurn:* Andrea Grêt, Jan Novak, Sandra Vitelli.
Local Principal Investigator: Frank-Peter Stephan

*Ente Ospedaliero Cantonale Bellinzona:* Jane Frangi-Kultalahti, Augusto Gallino, Luisa Vicari.
Local Principal Investigator: Marcello Di Valentino

*University of Zurich/University Hospital Zurich:* Helena Aebersold, Fabienne Foster, Matthias Schwenkglenks.

*Medical Image Analysis Center AG Basel:* Jens Würfel (Head), Anna Altermatt, Michael Amann, Marco Düring, Petra Huber, Esther Ruberte, Tim Sinnecker, Vanessa Zuber.

*Clinical Trial Unit Basel:* Michael Coslovsky (Head), Pascal Benkert, Gilles Dutilh, Milica Markovic, Pia Neuschwander, Patrick Simon

*Schiller AG Baar:* Ramun Schmid
